# The association between mitochondrial DNA copy number, low-density lipoprotein cholesterol, and cardiovascular disease risk

**DOI:** 10.1101/2022.10.23.22281418

**Authors:** Xue Liu, Xianbang Sun, Yuankai Zhang, Wenqing Jiang, Lai Meng, Kerri L. Wiggins, Laura M. Raffield, Lawrence F. Bielak, Wei Zhao, Achilleas Pitsillides, Jeffrey Haessler, Yinan Zheng, Thomas W. Blackwell, Jie Yao, Xiuqing Guo, Yong Qian, Bharat Thyagarajan, Nathan Pankratz, Stephen S. Rich, Kent D. Taylor, Patricia A. Peyser, Susan R. Heckbert, Sudha Seshadri, Eric Boerwinkle, Megan L. Grove, Nicholas B. Larson, Jennifer A. Smith, Ramachandran S. Vasan, Annette L. Fitzpatrick, Myriam Fornage, Jun Ding, April P. Carson, Goncalo Abecasis, Josée Dupuis, Alexander Reiner, Charles Kooperberg, Lifang Hou, Bruce M. Psaty, James G. Wilson, Daniel Levy, Jerome I. Rotter, Joshua C. Bis, TOPMed mtDNA Working Group in NHLBI Trans-Omics for Precision Medicine (TOPMed) Consortium, Claudia L. Satizabal, Dan E. Arking, Chunyu Liu

**Affiliations:** Department of Biostatistics, School of Public Health, Boston University, Boston, MA 02118, USA; Cardiovascular Health Research Unit, Department of Medicine, University of Washington, Seattle, WA 98101, USA; Department of Genetics, University of North Carolina at Chapel Hill, Chapel Hill, NC 27599, USA; Department of Epidemiology, School of Public Health, University of Michigan, Ann Arbor, MI 48109, USA; Fred Hutchinson Cancer Research Center, Division of Public Health Science, Seattle, WA 98109, USA; Feinberg School of Medicine, Northwestern University, Chicago, IL 60611, USA; TOPMed Informatics Research Center, University of Michigan, Ann Arbor, MI 48109, USA; The Institute for Translational Genomics and Population Sciences, Department of Pediatrics, The Lundquist Institute for Biomedical Innovation at Harbor-UCLA Medical Center, Torrance, CA 90502, USA; Longitudinal Studies Section, Translational Gerontology Branch, National Institute on Aging, NIH, Baltimore, MD 21224, USA; Department of Laboratory Medicine and Pathology, University of Minnesota, Minneapolis, MN 55455, USA; Department of Computational Pathology, University of Minnesota, Minneapolis, MN 55455, USA; Department of Public Health Services, Center for Public Health Genomics, University of Virginia, Charlottesville, VA 22908, USA; Cardiovascular Health Research Unit and Department of Epidemiology, University of Washington, Seattle, WA 98101, USA; Glenn Biggs Institute for Alzheimer’s and Neurodegenerative Diseases, University of Texas Health Science Center at San Antonio, San Antonio, TX 78229, USA; Framingham Heart Study, NHLBI, 73 Mt. Wayte Avenue, Suite 2, Framingham, MA 01702, USA; Department of Neurology, Boston University School of Medicine, Boston, MA 02118, USA; Human Genetics Center, Department of Epidemiology, Human Genetics and Environmental Sciences, The University of Texas Health Science Center at Houston, Houston, TX 77030, USA; Human Genome Sequencing Center, Baylor College of Medicine, Houston, TX, 77030, USA; Division of Clinical Trials and Biostatistics, Department of Quantitative Health Sciences, Mayo Clinic College of Medicine and Science, Rochester, MN 55905, USA; Sections of Preventive Medicine and Epidemiology, and Cardiovascular Medicine, Boston University School of Medicine, Boston, MA, 02118, USA; Departments of Family Medicine, Epidemiology, and Global Health, University of Washington, Seattle, WA 98195, USA; .Center for Human Genetics, University of Texas Health Science Center at Houston, Houston, TX 77030, USA; Department of Medicine, University of Mississippi Medical Center, Jackson, MS 39216, USA; Department of Epidemiology, Biostatistics and Occupational Health, School of Population and Global Health, McGill University Faculty of Medicine and Health Sciences, Montréal, QC H3G 2M1, Canada; Departments of Epidemiology, and Health Systems and Population Health, University of Washington, Seattle, WA 98101, USA; Department of Physiology and Biophysics, University of Mississippi Medical Center, Jackson, MS 39216, USA; Population Sciences Branch, National Heart, Lung, and Blood Institute, National Institutes of Health, Bethesda, MD 20892, USA; McKusick-Nathans Institute, Department of Genetic Medicine, Johns Hopkins University School of Medicine, Baltimore, Maryland, MD 21205, USA

## Abstract

Mitochondria are the primary organelle to generate cellular energy. Our group and others have reported that lower mitochondrial DNA copy number (mtDNA CN) is associated with higher risk of cardiovascular disease outcomes (CVD) and higher LDL levels. However, the causal relationship between mtDNA CN and CVD remains to be studied. Here we performed cross-sectional and prospective association analyses of blood-derived mtDNA CN and CVD outcomes in up to 27,316 participants from different racial/ethnic groups with whole genome sequencing. We validated most of the previously reported associations but effect sizes were smaller in this study. For example, one SD unit decrease in mtDNA CN was significantly associated with 1.08-fold (95% CI, 1.04, 1.12; *P*=1.7E-04) hazard for developing incident coronary heart disease (CHD) adjusting for age, sex and race/ethnicity. We conducted Mendelian randomization (MR) to explore causal relationships between mtDNA CN, LDL, and CHD. Bi-directional univariable MR analyses provided strong evidence indicating higher LDL level is causally associated with lower mtDNA CN, and CHD was weakly associated with lower mtDNA CN. We found no evidence supporting a causal association for lower mtDNA CN with higher CHD risk or higher LDL. In multivariable MR, no associations were observed between mtDNA CN and CHD controlling for LDL level (P =0.92), whereas strong evidence for a direct causal effect was found for higher LDL on lower mtDNA CN, adjusting for CHD status (P =8.3E-10). Findings from this study indicate high LDL underlies the complex relationships between vascular atherosclerosis and lower mtDNA CN.

## Introduction

Cardiovascular diseases (CVDs) are a leading cause of death globally.^1^ A large proportion of CVDs is due to atherosclerosis, an inflammation process in the blood vessels.^2,3^ Mitochondria are primary sites for oxidative phosphorylation machinery that generates adenosine triphosphate (ATP).^4^ Mitochondria have their own DNA (mtDNA), a circular 16.6-kb molecule encoding essential proteins for ATP production and energy homeostasis.^5^ The role of mtDNA integrity and mitochondrial dysfunction in the pathogenesis of atherosclerosis has been studied in genetic knockout mice with defects in the apolipoprotein E (ApoE) and the catalytic subunit of the mtDNA polymerase gamma subunit (POLG).^6,7^ In these knockout mice, mtDNA damage was an early event during the initiation of atherogenesis and may result from reactive oxygen species.^8^ Accumulation of mtDNA damage promoted atherosclerosis and was associated with the formation of vulnerable plaques.^9^ Moreover, trapping and oxidization of low-density lipoprotein (LDL) cholesterol at arterial walls also plays a central role in the formation of the atherosclerotic lesion and the progression of CVD.^3,10,11^ Accumulation in the subendothelial space of the arterial wall leads LDL to undergo oxidation to become oxidized LDL (oxLDL). This can lead to increased mitochondrial permeability, which may cause subsequent damage to mtDNA.^12,13^

Each human cell contains hundreds (e.g., in a blood cell) or even thousands (e.g., in a cardiac muscle cell) of mitochondria, and multiple copies of mtDNA are present per mitochondrion.^14^ The copy number of mtDNA (i.e., mtDNA CN) is strictly regulated for energy homeostasis and may serves as a surrogate marker of mitochondrial function.^15,16^ Thus, reduced mtDNA CN may serve as a biomarker of mitochondrial dysfunction.^17^ A lower level of mtDNA CN has also been associated with a general decline in health,^18^ all-cause mortality^18,19,20^, and associated with multiple cardiometabolic traits including higher level of LDL and hyperlipidemia that are major risk factors for CVD after controlling for other risk factors.^21,22^ Recent prospective studies have also reported significant associations between lower mtDNA levels and CVD outcomes.^19,23,24^ However, the causal relationship among mtDNA CN, LDL and CVD remains to be determined.

To that end, this study had two aims. The first aim was to validate the associations of mtDNA CN with CVD outcomes and total mortality using blood-derived mtDNA CN measured from whole genome sequencing (WGS) in eight cohorts of diverse populations. The previous mtDNA CN - CVD association studies used mtDNA CN measured by array-based methods or by qPCR in fewer cohorts.^19,23^ The second aim was to explore the causal relationships between mtDNA CN, LDL, and coronary heart disease (CHD) using Mendelian randomization (MR), a method that has been increasingly used to minimize issues of confounding and reverse causation with genetic variants as an instrumental variable **(Figure 1)**.^25^

**Figure 1.**
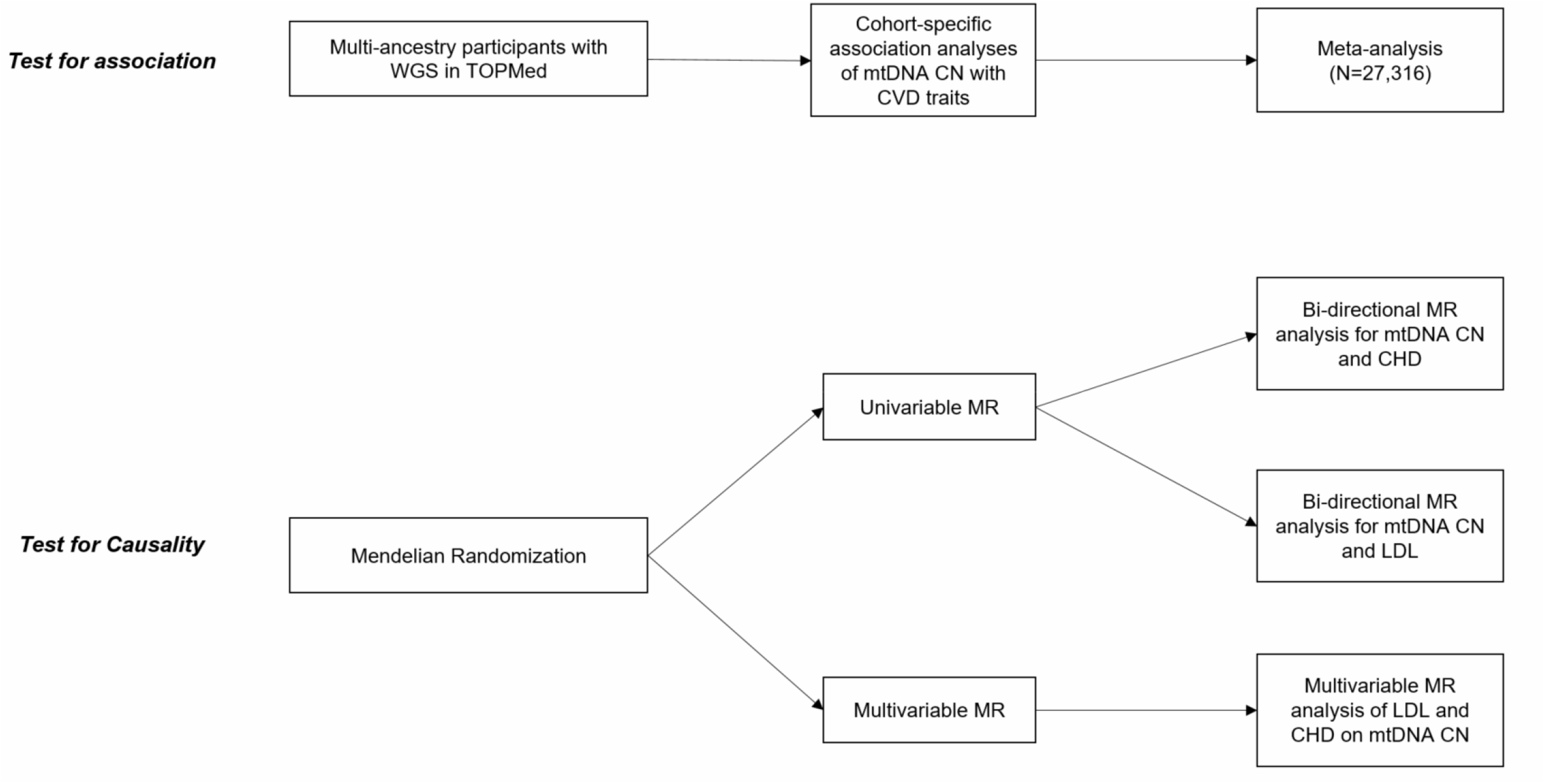
Study design. Association analysis of mtDNA cop number (CN) with cardiovascular disease traits was performed in eight cohorts of total 27,316 participants of multiple races/ethnicities. Meta-analysis was performed using the fixed effects inverse variance method to summarize the cohort specific results. Bi-directional univariable Mendelian Randomization was performed to test causality between mtDNA CN, CHD and LDL. Multivariable MR was performed to test the direct causal effect of LDL and CHD on mtDNA CN.

## Methods

### Study population

This study included participants with WGS from eight prospective cohort studies, some with participants from different ethnic/racial groups (**Supplemental Table 1**): the Atherosclerosis Risk in Communities study^26^ (ARIC) (n=3,585), the Coronary Artery Risk Development in Young Adults Study (CARDIA)^27^ (n=3,473), the Cardiovascular Health Study (CHS)^28^ (n=3,546), the Framingham Heart Study (FHS)^29,30,31^ (n=4,133), the Genetic Epidemiology Network of Arteriopathy (GENOA)^32^ (n=1,253), the Jackson Heart Study (JHS)^33^ (n=3,286), the Multi-Ethnic Study of Atherosclerosis Study (MESA)^34^ (n=4,596), and the Women’s Health initiative Study (WHI)^35^ (n=7,197). Except for WHI, in which only female participants were included, the other seven cohorts included both men and women. The CHS only included participants 65 and older (mean age 74 years) and the other cohorts included mostly middle-aged participants at blood-draw for this study (mean age ranging from 58 to 69). MESA excluded participants with any clinically recognized CVD at the baseline visit,^34^ while the other cohorts contained prevalent CVD cases at baseline. Several of the cohorts contained a small number of duplicate participants (n =136) due to study design and data collection^26,32,33^. We removed these duplicate participants from subsequent association analyses.

**Table 1.** Comparison of results between Multivariable and univariable Mendelian randomization.

| Exposure | N SNPs | Beta | Se | P |
| --- | --- | --- | --- | --- |
| Multivariable MR |  |  |  |  |
| CHD | 5 | -0.10 | 0.34 | 0.76 |
| LDL | 341 | 0.083 | 0.012 | 1.2E-12 |
| Univariable MR |  |  |  |  |
| CHD | 10 | -0.86 | 0.48 | 0.072 |
| LDL | 345 | 0.084 | 0.011 | 1.1E-14 |

Multivariable Mendelian randomization (MR) was performed for CHD and LDL on mtDNA CN. Univariable MR was performed for CHD on mtDNA CN, and LDL on mtDNA CN (IVW results were presented for univariable MR analyses). Q statistic for instrument strength of CHD is 1035, LDL is 3858. High heterogeneity was detected in multivariable MR IVW (Q value=1380, d.f.=340, $p = 4.5e-125$ )

### mtDNA CN estimation in whole genome sequencing

WGS was performed from one of the TOPMed sequencing centers using blood-derived DNA for all participants in the eight cohorts. The average genome-wide coverage was ∼39-fold across samples in TOPMed.^36^ The TOPMed Information Research Center conducted analyses to estimate mtDNA CN across all participants using the program *fastMitoCalc* of the software package *mitoAnalyzer*.^37^ Because nuclear DNA (nDNA) is diploid while mitochondrial DNA is haploid, the average mtDNA CN per cell was estimated as twice the ratio of the average coverage of mtDNA to the average coverage of the nuclear DNA (nDNA).^37^

### Cardiovascular disease traits and total mortality

The eight longitudinal cohorts in this study have been established to investigate risk factors contributing to CVD, morbidity and mortality. Each cohort used standardized definitions to adjudicate CVD outcomes. CHD was defined as the first incident myocardial infarction (MI) or death owing to CHD and cardiac procedures (typically revascularization).^38^ Stroke was defined as the first nonfatal stroke or death owing to stroke.^39^ Heart failure is a complex clinical syndrome resulting from a structural or functional cardiac disorder that impairs the ability of one or both ventricles to fill with or eject blood sufficiently to meet the needs of the body.^40,41^ CVD included CHD, stroke, and heart failure, and death due to CHD, stroke and heart failure. All-cause mortality included all deaths. We analyzed associations of mtDNA CN with three prevalent and incident CVD outcomes (CHD, stroke, and CVD) and with all-cause mortality.

### Covariates

In the primary analysis, age at blood draw, sex, study center (if applicable), and self-reported racial/ethnic group were adjusted for in the base model. Additional variables included body mass index (BMI, kg/m^2^), fasting plasma lipid measures including total cholesterol (TC, mg/dL) and high density lipoprotein cholesterol (HDL, mg/dL), systolic blood pressure (SBP, mmHg), treatment for high blood pressure or hypertension (HRX), current smoking status, and diabetes status. Diabetes was defined as fasting blood glucose level of ≥126 mg/dL or currently receiving medications to lower blood glucose levels to treat diabetes. This study used mtDNA CN calculated using WGS of blood-derived DNA. Different blood cell types (e.g. neutrophils, lymphocytes, etc…) contain different levels of mtDNA CN.^42,43^ To minimize potential confounding, we accounted for white blood cell count and differential components (the proportions of neutrophils, lymphocytes, monocytes, eosinophils, and basophils) and platelet count in association analyses in cohorts in which these cell count variables were available.^21^

### Association analyses of mtDNA CN with CVD outcomes and total mortality

For primary analyses, we generated mtDNA CN residuals by regressing mtDNA CN on age, age-squared, sex and blood collection year (as a categorical variable) in each cohort.^21^ For age-stratified analysis, we generated mtDNA CN residuals by regressing mtDNA on sex and blood collection year in each cohort. For sex-stratified analysis, we generated mtDNA CN residuals by regressing mtDNA on age, age-squared, and blood collection year in each cohort. The residuals were standardized to a mean of zero and standard deviation (s.d.) of one. The standardized residuals were used as the main predictor in all regression models.^21^ We removed participants whose mtDNA CN standardized residuals were greater than 4 s.d. from the mean.

We performed cohort-specific association analyses between mtDNA CN and outcomes. We used logistic regression to quantify the associations of mtDNA CN with prevalent CVD outcomes. We used a Cox proportional hazards regression model to quantify the association of mtDNA CN with incident CVD outcomes and total mortality in all cohort-specific analyses. Due to a special study design in selecting participants for WGS in WHI, we applied a weighted logistic regression for cross-sectional outcome or a weighted Cox proportional hazards regression for incident outcomes in WHI (**Supplemental Methods**). We performed three models for association analyses of mtDNA CN with both prevalent and incident outcomes. Model 1 included age, sex, study center (if applicable), and race/ethnicity. In Model 2, we additionally adjusted for several traditional covariates including BMI, TC, HDL, SBP, HRX, current smoking and diabetes for CVD outcomes. For analyzing total mortality as the outcome, we excluded participants who have prevalent CHD or diabetes and adjusted for BMI, TC, HDL, SBP, HRX and current smoking^19^ in Model 2. In Model 3, white blood cell and differential counts as well as platelet counts were further adjusted in addition to covariates in Model 2. We used an inverse variance meta-analysis with a fixed effects model to summarize cohort-specific association analyses. An odds ratio (OR) or a hazard ratio (HR) was reported corresponding to one s.d. decrease in the mtDNA CN level.

In secondary analyses, we performed association analyses between mtDNA CN and outcomes in: 1) male and female only samples, and 2) in participants who were at least aged 60 years at blood draw for WGS. We also performed several sensitivity analyses in FHS to investigate if different cardiometabolic disease status (hypertension, diabetes and hyperlipidemia) may result in different directionalities or effect sizes in associations of mtDNA CN with CVD.

### Mendelian randomization

LDL plays a central role in the pathogenesis of CVD outcomes including CHD. To thoroughly evaluate the causal relationship between CHD and mtDNA CN, we first conducted univariable bi-directional two sample MR analysis^44^ to assess the causal relationship between LDL and mtDNA CN, and between CHD and mtDNA CN. We used single nucleotide polymorphisms (SNPs) (LD *r*^2^< 0.001 based on EUR population reference panel) with no ambiguous allele information (i.e., palindromic SNPs).^44^ We further excluded SNPs that are known to be pleiotropic in MR analysis (i.e., the missense mutations rs7412, rs429358, rs768374191 and rs367866106 in ApoE).^45^ We used the inverse variance weighted (IVW) method to combine the causal effects of independent SNPs. Leave-one-out plots were used to detect influential outliers and MR-PRESSO was used to detect and correct for potential outliers.^46^ We also conducted several sensitivity analyses, including MR-Egger regression, Cochran’s Q statistic and funnel plots, and obtained median and mode estimates to test the validity of MR estimators.^47,48,49,50,51^ In secondary analysis to test causality of mtDNA CN to CHD, we conducted MR analysis using SNPs identified by Gene Ontology analysis. These selected SNPs are directly involved in mitochondrial functions. (**Supplemental Table 2**).^52^ TwoSampleMR package (version 0.5.0) in R (version 0.5.6) was used for univariable MR analyses.

Because common genetic variants are associated with both LDL and CHD, horizontal pleiotropy effect may exist, which violates the 3rd assumption of MR.^49,53,54^ To account for this potential pleiotropic effect and to simultaneously investigate the direct effect of either LDL or CHD on mtDNA CN, we conducted multivariable MR (MVMR).^55,56,57^ We performed the extended framework of IVW and MR-Egger methods to estimate causal effects in multivariable MR analysis.^58,59^ We used GWAS results from non-overlapping participants for each exposure (see below) and thus the covariance between the effects of the genetic variants on each exposure was fixed at zero. We used generalized Cochran’s Q test to assess instrumental variable (IV) validity in the two-sample summary data setting.^55,60^ The MVMR package (version 0.2.0) in R (version 0.5.6) was used for MVMR analyses.

In all MR analyses, we used significant SNPs (*p* < 5e-8) as IVs identified from genome-wide association studies (GWAS). The significant SNPs of mtDNA CN GWAS were recently reported from the Cohorts of Heart and Aging Research in Genomic Epidemiology (CHARGE) consortium and the UK Biobank data.^61^ In the present study, we used the mtDNA CN GWAS results for the selected SNPs from the ∼300,000 UK Biobank only participants analysis.^61^ We identified the CHD-associated SNPs list and extracted the summary GWAS results for the identified SNPs from UK Biobank participants (10,157 cases and 351,037 controls, 2018, the Neale lab, https://gwas.mrcieu.ac.uk/datasets/ukb-d-I9_CHD/). We also identified SNPs from the Global Lipids Genetics Consortium summary GWAS results of LDL from analyses of 1,166,583 multi-ancestries participants (Graham, Kanoni, Ramdas, et al., 2021 http://csg.sph.umich.edu/willer/public/glgc-lipids2021/).

## Results

### Participant characteristics

This study included up to 27,316 participants (mean age 62, age range of 20-98 years, and 68% women) with 16636 (60.9%) European Americans (EA), 8709 (31.9%) African Americans (AA), 1229 (4.5%) Hispanic/Latino Americans (HA) and 728 (2.7%) East Asian Americans (**Supplemental Table 1**) (EAS). The prevalence of any CVD outcomes was higher in African Americans (13.4%) than other ethnic and racial groups (EA: 9.4%, HA: 9.8%, EAS: 10.2%). During a median of 6 to 16 years (across the cohorts) of follow-up, the prevalence and incidence rates of CVD outcomes varied across cohorts (**Supplemental Table 1**).

### Association analyses of mtDNA CN with prevalent CVD outcomes

In total, 2,162 (7.9%) participants had prevalent CHD, 1154 (4.2%) had prevalent stroke, and 3397 (12.4%) had prevalent CVD at baseline (**Supplemental Table 1**). Meta-analysis showed that one s.d. decrease in mtDNA CN was significantly associated with a 1.11-fold odds of CHD (95% CI, 1.07, 1.16; *P*=1.3E-07), 1.13-fold odds of stroke (95% CI, 1.05, 1.22; *P*=0.002) and 1.14-fold odds of CVD (95% CI, 1.11, 1.16; P=1.9E-33), adjusting for age, sex and race/ethnicity (**Figure 2**, Model 1). The associations were slightly attenuated after further adjusting for traditional CVD risk factors (**Supplemental Figure 1**, Model 2) and white blood cell count in addition to traditional CVD risk factors (**Supplemental Figure 2**, Model 3). The association directions were consistent across 6 of the 7 cohorts with one null association for CVD outcomes (**Figure 2**).

**Figure 2.**
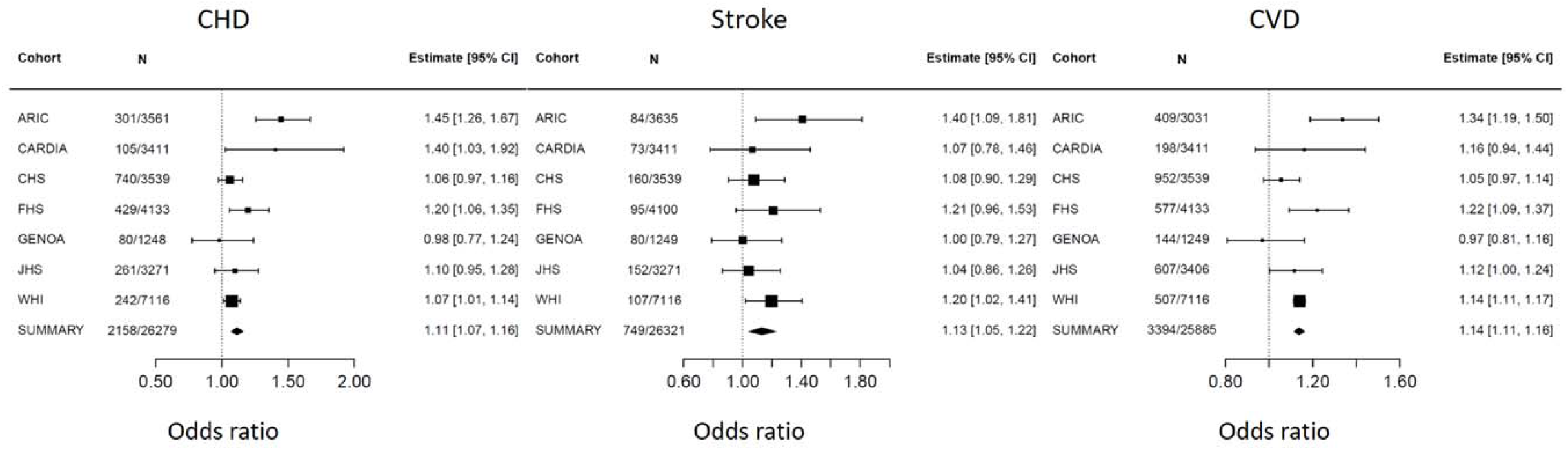
Association and meta-analysis of mtDNA CN and prevalent CVD outcomes. We performed logistic regression with an outcome and mtDNA residuals as independent variable adjusting for age, sex, study center (if applicable), and race/ethnicity in Model1. The size of the square represent the weight of each cohort. ARIC, Atherosclerosis Risk in Communities study; CARDIA, Coronary Artery Risk Development in Young Adults Study; CHS, Cardiovascular Health Study; FHS, Framingham Heart Study; GENOA, Genetic Epidemiology Network of Arteriopathy Study; JHS, Jackson Heart Study; MESA, Multi-Ethnic Study of Atherosclerosis; WHI, Women’s Health initiative

### Association analyses of incident CVD outcomes and all-cause mortality

A total of 24,019 participants free of CVD at baseline was followed up for a median of 12 years. During the follow-up, 3975 (16.5%) had incident CHD, 5208 (21.7%) had incident stroke, and 8590 (35.4%) had incident CVD. Meta-analysis showed that one s.d. decrease in mtDNA CN was significantly associated with a 1.08-fold of hazards for developing incident CHD (95% CI, 1.04, 1.12; *P*=1.7E-04), 1.07-fold of hazards for developing incident CVD (95% CI, 1.03, 1.10; *P*=3.9E-05) when we adjusted for age, sex and race/ethnicity in association analyses **(Figure 3)**. The associations were slightly attenuated after further adjusting for traditional CVD risk factors in Model 2 (Incident CHD: HR =1.05; 95% CI, 1.01, 1.09; *P*= 0.023; Incident CVD: HR =1.05; 95% CI, 1.02, 1.09; *P*= 2.3E-03) (**Supplemental Figure 3**). The associations were also changed little after additionally adjusting for white blood cell count/differential count and platelet count in Model 3 (Incident CHD: HR =1.07; 95% CI, 1.02, 1.12; *P*= 4.3E-03; Incident CVD: HR =1.06; 95% CI, 1.03, 1.10; *P*= 9E-04) (**Supplemental Figure 4**). Incident stroke was not significantly associated with mtDNA CN in meta-analyses of the three models (**Supplemental Figure 3, Supplemental Figure 4**).

**Figure 3.**
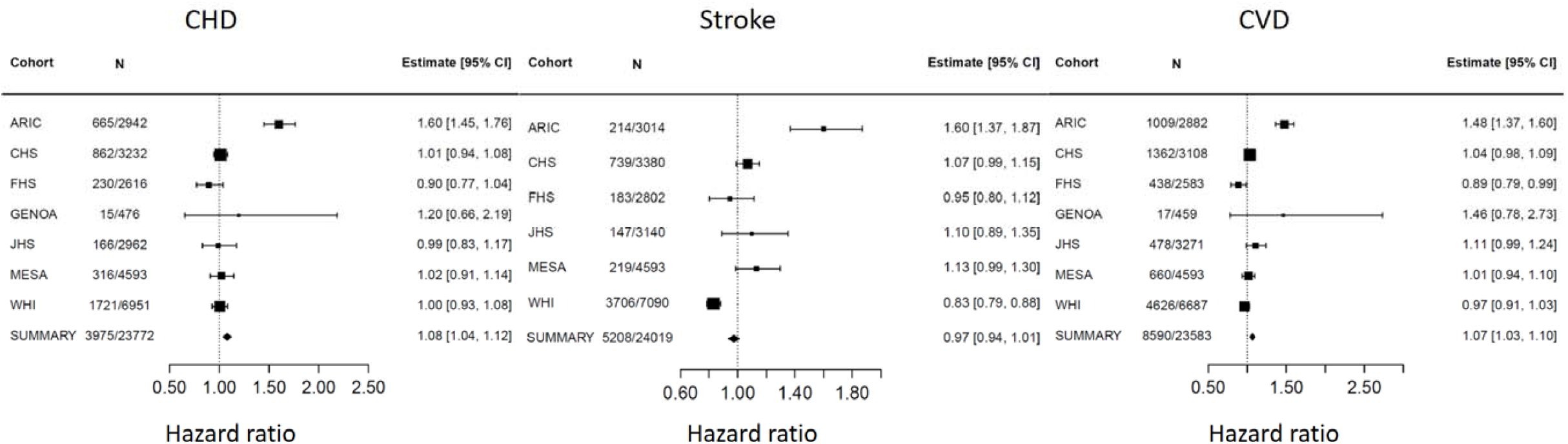
Association and meta-analysis of mtDNA CN and incident CVD outcomes. We performed Cox proportional hazards regression with an outcome and mtDNA residuals as independent variable adjusting for age, sex, study center (if applicable), and race/ethnicity in Model 1. The size of the square represent the weight of each cohort. ARIC, Atherosclerosis Risk in Communities study; CARDIA, Coronary Artery Risk Development in Young Adults Study; CHS, Cardiovascular Health Study; FHS, Framingham Heart Study; GENOA, Genetic Epidemiology Network of Arteriopathy Study; JHS, Jackson Heart Study; MESA, Multi-Ethnic Study of Atherosclerosis; WHI, Women’s Health initiative

Examining the individual cohorts, we found that lower mtDNA CN was associated with higher hazards for developing incident CHD and incident CVD in five cohorts, with ARIC displaying the strongest associations while FHS and WHI showed weak inverse associations or no association (**Figure 3**). Sensitivity analysis removing ARIC showed non-statistically significant result. Additional sensitivity analyses in FHS demonstrated that several factors, including age, sex, hypertension status, diabetes status and hyperlipidemia status were not the cause for the inverse effect or null association observed in FHS compared to other cohorts although the magnitude of associations seemed different in stratified analyses (**Supplemental Table 3**).

A total of 8018 (33.3%) participants died due to any cause during the follow-up. One s.d. decrease in mtDNA CN was significantly associated with 1.06-fold of hazards for all-cause mortality (95% CI, 1.03-1.09; *P*=2.5E-05) adjusting for age, sex and race/ethnicity. All the cohorts showed consistent directionality between mtDNA CN and total mortality in Model 1, that is, lower mtDNA CN was associated with higher rates of all-cause mortality (**Figure 4**). The associations were very similar after further adjusting for multiple clinical covariates in Model 2 (HR =1.05, 95% CI, 1.02-1.08; *P*= 5.9E-04) and additionally adjusting for cell counts/differential components and platelet count in Model 3 (HR =1.06, 95% CI, 1.02-1.10; *P*= 1.1E-03) (**Supplemental Figure 5**).

**Figure 4.**
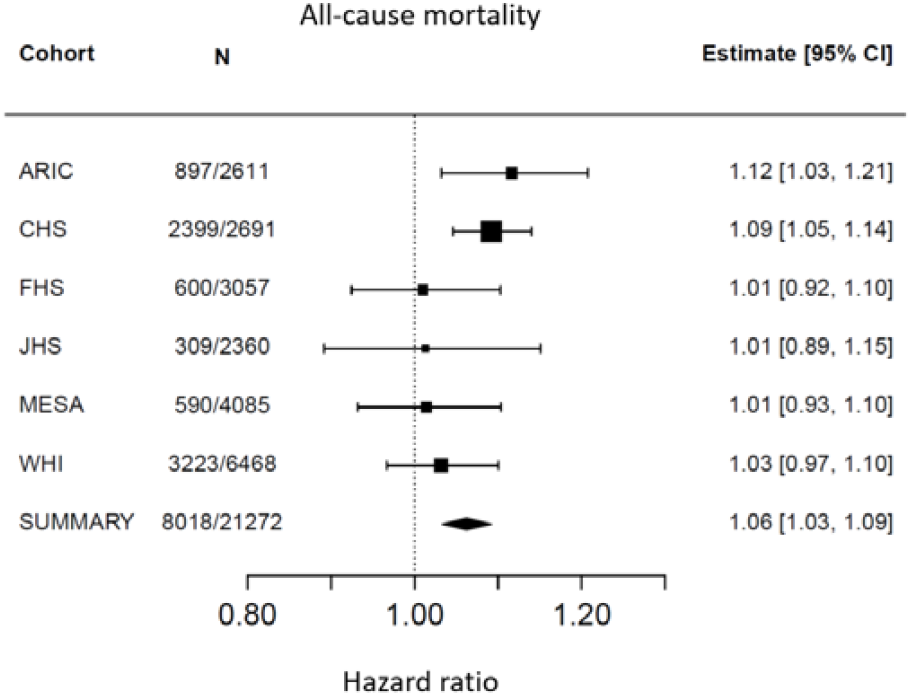
Association and meta-analysis of mtDNA CN and all-cause mortality. We performed Cox proportional hazards regression with an outcome and mtDNA residuals as independent variable adjusting for age, sex, study center (if applicable), and race/ethnicity in Model 1. The size of the square represent the weight of each cohort. ARIC, Atherosclerosis Risk in Communities study; CARDIA, Coronary Artery Risk Development in Young Adults Study; CHS, Cardiovascular Health Study; FHS, Framingham Heart Study; GENOA, Genetic Epidemiology Network of Arteriopathy Study; JHS, Jackson Heart Study; MESA, Multi-Ethnic Study of Atherosclerosis; WHI, Women’s Health initiative

### Mendelian randomization analyses to test causality

We performed bi-directional univariate MR analyses to test the causal relationships between mtDNA CN, LCL, and CHD. We used 77 (primary analysis) independent SNPs (LD *r*^*2*^ < 0.001) as IVs to test the causal relationship of mtDNA CN on CHD (**Supplemental Table 4**). The IVW MR analyses yielded insufficient evidence (OR = 1.0, 95% CI, 0.997, 1.010; *P*=0.25) to support that lower mtDNA CN was casually associated with higher odds of CHD. Sensitivity analyses and the secondary analysis with 23 SNPs that are directly involved in mitochondrial function further confirmed no causal relationship from mtDNA CN to CHD (**Supplemental Table 5, Supplemental Figure 6)**. To test for the causal relationship of CHD on mtDNA CN, we used 10 significant GWAS SNPs (LD *r*^*2*^ < 0.001) (**Supplemental Table 7**) as IVs in MR analyses. The IVW MR analysis supported that having CHD displayed a borderline significant causal relationship with a lower mtDNA CN level (beta = -0.86; 95% CI, - 1.80, -0.08; *P*=0.072; Egger *P*=0.067) (**Supplemental Table 6, Supplemental Figure 7**). For all the above MR analyses, heterogeneity was observed although results remained unchanged after correction by MR-PRESSO. In addition, the MR-Egger test did not provide significant evidence of directional pleiotropy **(Supplemental Table 5)**. To account for the observed heterogeneity with the 10 SNPs (*P* = 9.6E-09), we applied multiple sensitivity analyses to test for validity of MR estimators. Three of six sensitivity analyses gave rise to significant causal effects of CHD on mtDNA CN (**Supplemental Table 6**). For example, weighted median test (beta = -1.06; *P* = 0.0036) and weighted mode (beta = -1.48; *P* = 0.0041).

We selected 75 independent GWAS SNPs associated with mtDNA CN (**Supplemental Table 8**) to test the causal relationship of mtDNA CN on LDL. The IVW MR analysis gave rise to weak evidence that lower mtDNA CN was borderline significantly associated with a higher LDL (beta=0.067, 95% CI, -0.0036, 0.14; *P*=0.059; Egger P=0.22) **(Supplemental Table 9, Supplemental Figure 8)**. The corrected effect of mtDNA CN on LDL is not statistically significant (beta=0.025, 95% CI= -0.0024, 0.052, P-value= 0.08) after removing outliers identified by the MR-PRESSO test. No directional pleiotropy was detected using the MR-Egger method (**Supplemental Table 9**). All sensitivity analyses yielded non-statistically significant causal effect of mtDNA CN on LDL-C (*P* > 0.05, **Supplemental Table 9**).

To test the causal relationship of LDL-C on mtDNA CN, 345 SNPs were selected as IVs (**Supplemental Table 10)**. The IVW MR analysis showed a strong causal relationship between lower LDL and higher mtDNA CN (beta=0.084, 95% CI=0.062, 0.11, P-value=1.1E-14). High heterogeneity was observed in the IVW analysis and thus MR-PRESSO was performed. After outliers were removed by MR-PRESSO, the corrected causal effect became more statistically significant (beta=0.077, 95% CI=0.062, 0.092, P-value=1.0E-21) than the IVW MR result. All the sensitivity analyses presented statistically significant causal relationship of LDL on mtDNA CN (P<0.0001) (**Supplemental Table 9**).

Given the findings from univariable MR analyses and the previous findings that LDL is a primary cause for CHD development,^10,11^ we conduced MVMR to estimate the direct effect of either CHD or LDL on mtDNA CN controlling for each other. After excluding SNPs with a pairwise >0.001, 346 SNPs were used in the analysis. We observed strong evidence for a direct causal effect of LDL on mtDNA CN adjusting for CHD, that is, one s.d. decrease in genetically predicted LDL level was causally associated with 0.083 s.d. higher mtDNA CN level (IVW beta=0.083, 95% CI= 0.059, 0.11, *P* =1.2E-12) (**Table 1, Figure 4**). In contrast, the direct causal effect of CHD on mtDNA CN was not significant controlling for LDL level (IVW beta=-0.1, 95% CI= -0.57, 0.77, *P* =0.76) (**Table 1**). The MVMR-Egger test yielded consistent results as those from IVW MVMR analysis for both LDL and CHD.

## Discussion

In this study, we validated the association of mtDNA CN with prevalent and incident CVD outcomes (except for incident stroke), as well as all-cause mortality during a median 12 years of follow-up in up to 27,316 participants from eight cohort studies including self-identified European Americans, African Americans, Hispanic/Latino Americans, and East Asian Americans. The associations of mtDNA CN with the outcome variable remained statistically significant after further adjustment for traditional clinical variables (i.e. total cholesterol and HDL, etc) and blood cell counts. More importantly, we performed comprehensive univariable and multivariable MR analyses, using SNPs identified from the latest GWAS for CHD^44,62,63,64^, LDL^65,66^ and mtDNA CN^61^ to explore the causal relationships between mtDNA CN, LDL and CHD. We applied the MR-PRESSO and MR-Egger tests as well as several sensitivity analyses to minimize the bias and to test validity of MR analyses. The MR analyses implicate that elevated LDL levels as the primary driver for the observed significant association of mtDNA CN with CHD.

It has always been challenging to assess causality in epidemiological association analyses. The bi-directional univariable MR analyses found that having CHD seemed to have a causal effect on lower mtDNA CN level rather than an opposite direction that lower mtDNA CN had causal effect on CHD. CHD is a multifactorial end-point disease that is characterized as the reduction of blood flow to the heart muscle due to build-up of atherosclerotic plaque.^3^ Studies have consistently shown that atherosclerosis is initiated by excess LDL levels in the plasma. In addition, our recent study reported that higher LDL levels are associated with lower mtDNA CN in blood.^21^ Thus, it was necessary to consider LDL-possibly play a major role in assessing the causal relationship between CHD and mtDNA CN. Bi-directional MR supported that LDL has a causal effect on mtDNA CN while mtDNA CN had no causal effect on LDL. Given it is well known that LDL is a causal factor for the development of CHD^67^ and that LDL and CHD share common genetic variants, we performed a MVMR analysis to assess the direct causal effect of CHD or LDL on mtDNA CN. We observed a significant, direct causal effect of LDL on mtDNA CN adjusting for CHD (**Figure 5A**). In contrast, the direct causal effect of CHD on mtDNA CN became insignificant controlling for LDL. Based on these findings, it is reasonable to conclude that the observed association between mtDNA CN and CVD outcomes (prevalent and incident) is a manifestation of the causal effect of higher LDL levels on lower mtDNA CN.

**Figure 5.**
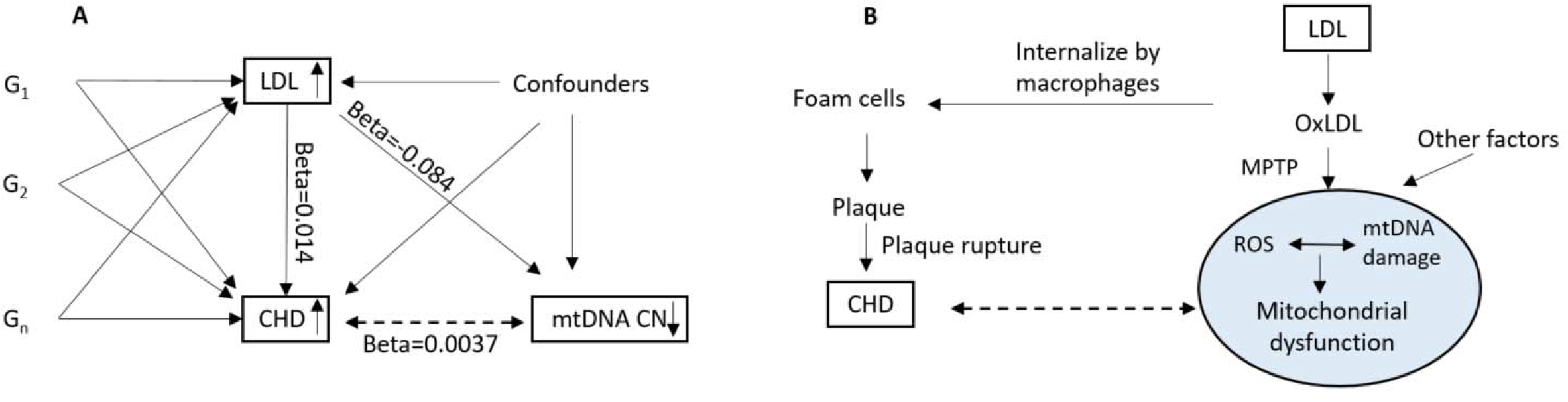
The Causal relationships between LDL, CHD and mtDNA CN. A. Mendelian randomization. B. Biological experiment (adapted from Lee at al.^3^). LDL: LDL cholesterol. CHD: coronary heart disease. mtDNA CN: mitochondrial DNA copy number. OxLDL: oxidized LDL. MPTP: mitochondrial permeability transition pore. ROS: reactive oxygen species. Univariable MR analysis demonstrated that one s.d. lower in genetically predicted LDL was causally associated with 0.99 hazard ratio of CHD. Multivariable MR analysis demonstrated the direct effect of one s.d. lower in genetically predicted LDL was causally associated with 0.092 s.d. lower in mtDNA CN (P=3.2E-09), and the direct effect of genetically predicted CHD condition was not associated with mtDNA CN (beta= -0.039, P=0.92).

Our findings are supported by recent advances focusing on elucidating the role of oxidative stress and mitochondrial dysfunction in vascular inflammation and atherosclerosis in animal models.^6,7,8^ LDL in the plasma is the primary molecule that triggers a cascade of inflammation responses. The excess of LDL is oxidized into oxidized LDL (oxLDL), which attracts immune cells like monocytes into the arterial wall. Monocytes then differentiate into macrophages which swallow oxLDL and become foam cells, the most abundant immune cells within the atherosclerotic lesion.^6,7,8^ CHD occurs when the plague buildup is ruptured to form a large thrombus. On the other hand, the oxLDL and other factors can increase the permeability of the mitochondrial transition pore, lead to the swelling of the mitochondrial matrix, and the increase in ROS production in mitochondria.^68,69^ The elevated ROS production in mitochondria may damage the mtDNA and eventually result in mitochondrial dysfuntion.^70^ The MR analyses in our study support that higher LDL is the driver for CHD and lower mtDNA CN levels (**Figure 5B**). Nonetheless, the role of mtDNA CN in the atherosclerotic formation, the pathogenesis of CVD, and inflammation is complex and warrants further investigation.

### Limitations of the study

Heterogeneity was observed in the association estimates of CVD outcomes across cohorts even though we harmonized phenotypes and accounted for confounders and known batch effects in association analyses with mtDNA CN. This remaining heterogeneity may partially be related to different distributions in age, sex and CVD phenotypes across study cohorts. Experiment conditions for blood draws, DNA extraction, storage and other unobserved confounding factors may also have contributed to the heterogeneity. For the MVMR analysis, we only included LDL which is known to associate with mtDNA CN and play a causal role in CHD to investigate the direct causal effects of LDL and CHD on mtDNA CN. Other risk factors, e.g., triglycerides and diabetes, were not considered in MVMR. Therefore, horizontal pleiotropy was not fully accounted for, which was also reflected by a large test statistic in testing heterogeneity in MVMR **(Table 1)**. However, MR-PRESSO is currently not available to correct for pleiotropic effects in MVMR.

### Strength of the study

The main strength of this study is that we adopted bi-directional and multivariable MR analysis to disentangle the complex relationship between mtDNA CN and CHD in cohort studies. Observational epidemiological studies are susceptible to confounding and subclinical disease stage may impact observed associations between CVD outcomes and mtDNA CN.^71^ Robust genetic variants have been identified in large GWAS with mtDNA CN (n = 465,809), CHD (n = 361,194) and LDL (n=1,166,583).^61,62,72,73,74^ To minimize bias in MR analyses, we removed known pleiotropic SNPs (e.g., *APOE* SNPs) that are associated with both mtDNA CN and CVD traits. We performed MR IVW as well as sensitivity analyses including MR-Egger, Median and Mode methods to provide evidence for validity of MR estimators. Benefiting from the widely available GWAS data, we were able to utilize non-overlapping samples for GWAS of each exposure, which strengthened the validity of the two-sample MVMR analysis.^60^ Overall, the multivariable MR results provide evidence that higher LDL level is the driving factor for the association between lower mtDNA CN and CHD. An additional strength is that, in most of these cohorts, the CVD outcomes and all-cause death data, have been regularly collected for adjudicated by a physician endpoints review committee.^75,76^ The well-characterized outcome and predictor variables and hierarchical association analyses with three models to reduce potential confounding.

In summary, this study validated the previously reported association of mtDNA CN with CVD outcomes and all-cause mortality. In addition, we used both univariable and multivariable MR analyses to demonstrate an independent causal effect of LDL underlying both mtDNA CN and CHD, even after accounting for other risk factors. Findings from this study add to an increasing volume of evidence surrounding the harmful effects of high LDL in the complex relationships between vascular inflammation, atherosclerosis, lower mtDNA CN, and mitochondrial dysfunction. Therefore, control for LDL and inflammation may be a feasible therapeutic strategy to improve mitochondrial function and cardiovascular health.

## Data Availability

All data produced are available online at TOPMed

## Acknowledgments

We included detailed acknowledgment for each cohort in Supplemental Materials. In brief, the authors thank the staff and participants of the Atherosclerosis Risk in Communities study, Cardiovascular Health Study, Coronary Artery Risk Development in Young Adults Study, Framingham Heart Study, Jackson Heart Study, Genetic Epidemiology Network of Arteriopathy Study, Multi-Ethnic Study of Atherosclerosis Study, and Women’s Health Initiative. We gratefully acknowledge the studies and participants who provided biological samples and data for TOPMed. Whole genome sequencing (WGS) for the Trans-Omics in Precision Medicine (TOPMed) program was supported by the National Heart, Lung and Blood Institute (NHLBI). Centralized read mapping and genotype calling, along with variant quality metrics and filtering were provided by the TOPMed Informatics Research Center (R01HL-117626-02S1; contract HHSN268201800002I). Phenotype harmonization, data management, sample-identity QC, and general study coordination were provided by the TOPMed Data Coordinating Center (R01HL-120393-02S1; contract HHSN268201800001I). Additional phenotype harmonization was performed by the current study (R01AG059727). The views expressed in this manuscript are those of the authors and do not necessarily represent the views of the National Heart, Lung, and Blood Institute; the National Institutes of Health; or the U.S. Department of Health and Human Services.

## References

1. Organization WH. Cardiovascular diseases. 2020.

2. Willerson JT, Ridker PM. Inflammation as a Cardiovascular Risk Factor. Circulation 2004;109:II-2-II-10.

3. Lee YT, Lin HY, Chan YWF, et al. Mouse models of atherosclerosis: a historical perspective and recent advances. Lipids in Health and Disease 2017;16:12.

4. Doenst T, Nguyen TD, Abel ED. Cardiac metabolism in heart failure: implications beyond ATP production. Circ Res 2013;113:709–24.

5. Taylor RW, Turnbull DM. Mitochondrial DNA mutations in human disease. Nature Reviews Genetics 2005;6:389–402.

6. Trifunovic A, Wredenberg A, Falkenberg M, et al. Premature ageing in mice expressing defective mitochondrial DNA polymerase. Nature 2004;429:417–23.

7. Kujoth GC, Hiona A, Pugh TD, et al. Mitochondrial DNA mutations, oxidative stress, and apoptosis in mammalian aging. Science 2005;309:481–4.

8. Ballinger SW, Patterson C, Knight-Lozano CA, et al. Mitochondrial integrity and function in atherogenesis. Circulation 2002;106:544–9.

9. Yu E, Baker L, Harrison J, et al. Mitochondrial DNA damage promotes atherosclerosis and is associated with vulnerable plaque. The Lancet 2013;381:S117.

10. Alfaddagh A, Martin SS, Leucker TM, et al. Inflammation and cardiovascular disease: From mechanisms to therapeutics. Am J Prev Cardiol 2020;4:100130.

11. Kobiyama K, Ley K. Atherosclerosis. Circulation research 2018;123:1118–20.

12. Di Lisa F, Canton M, Menabo R, Dodoni G, Bernardi P. Mitochondria and reperfusion injury. The role of permeability transition. Basic Res Cardiol 2003;98:235–41.

13. Halestrap AP, Clarke SJ, Javadov SA. Mitochondrial permeability transition pore opening during myocardial reperfusion--a target for cardioprotection. Cardiovasc Res 2004;61:372–85.

14. Miller FJ, Rosenfeldt FL, Zhang C, Linnane AW, Nagley P. Precise determination of mitochondrial DNA copy number in human skeletal and cardiac muscle by a PCR-based assay: lack of change of copy number with age. Nucleic Acids Res 2003;31:e61.

15. Reznik E, Miller ML, Senbabaoglu Y, et al. Mitochondrial DNA copy number variation across human cancers. Elife 2016;5.

16. Malik AN, Czajka A. Is mitochondrial DNA content a potential biomarker of mitochondrial dysfunction? Mitochondrion 2013;13:481–92.

17. Castellani CA, Longchamps RJ, Sun J, Guallar E, Arking DE. Thinking outside the nucleus: Mitochondrial DNA copy number in health and disease. Mitochondrion 2020;53:214–23.

18. Mengel-From J, Thinggaard M, Dalgard C, Kyvik KO, Christensen K, Christiansen L. Mitochondrial DNA copy number in peripheral blood cells declines with age and is associated with general health among elderly. Hum Genet 2014;133:1149–59.

19. Ashar FN, Moes A, Moore AZ, et al. Association of mitochondrial DNA levels with frailty and all-cause mortality. J Mol Med (Berl) 2015;93:177–86.

20. Fazzini F, Lamina C, Fendt L, et al. Mitochondrial DNA copy number is associated with mortality and infections in a large cohort of patients with chronic kidney disease. Kidney Int 2019;96:480–8.

21. Liu X, Longchamps RJ, Wiggins KL, et al. Association of mitochondrial DNA copy number with cardiometabolic diseases. Cell Genomics 2021;1:100006.

22. Tin A, Grams ME, Ashar FN, et al. Association between Mitochondrial DNA Copy Number in Peripheral Blood and Incident CKD in the Atherosclerosis Risk in Communities Study. J Am Soc Nephrol 2016;27:2467–73.

23. Ashar FN, Zhang Y, Longchamps RJ, et al. Association of Mitochondrial DNA Copy Number With Cardiovascular Disease. JAMA Cardiol 2017;2:1247–55.

24. Zhang Y, Guallar E, Ashar FN, et al. Association between mitochondrial DNA copy number and sudden cardiac death: findings from the Atherosclerosis Risk in Communities study (ARIC). European Heart Journal 2017;38:3443–8.

25. Burgess S, Foley CN, Allara E, Staley JR, Howson JMM. A robust and efficient method for Mendelian randomization with hundreds of genetic variants. Nat Commun 2020;11:376.

26. The Atherosclerosis Risk in Communities (ARIC) Study: design and objectives. The ARIC investigators. Am J Epidemiol 1989;129:687–702.

27. Friedman GD, Tekawa I, Grimm RH, Manolio T, Shannon SG, Sidney S. The Leucocyte Count: Correlates and Relationship to Coronary Risk Factors: The CARDIA Study. International Journal of Epidemiology 1990;19:889–93.

28. Fried LP, Borhani NO, Enright P, et al. The Cardiovascular Health Study: design and rationale. Ann Epidemiol 1991;1:263–76.

29. Dawber TR, Meadors GF, Moore FE, Jr. Epidemiological approaches to heart disease: the Framingham Study. Am J Public Health Nations Health 1951;41:279–81.

30. Feinleib M, Kannel WB, Garrison RJ, McNamara PM, Castelli WP. The Framingham Offspring Study. Design and preliminary data. Prev Med 1975;4:518–25.

31. Splansky GL, Corey D, Yang Q, et al. The Third Generation Cohort of the National Heart, Lung, and Blood Institute’s Framingham Heart Study: design, recruitment, and initial examination. Am J Epidemiol 2007;165:1328–35.

32. Daniels PR, Kardia SL, Hanis CL, et al. Familial aggregation of hypertension treatment and control in the Genetic Epidemiology Network of Arteriopathy (GENOA) study. Am J Med 2004;116:676–81.

33. Wilson JG, Rotimi CN, Ekunwe L, et al. Study design for genetic analysis in the Jackson Heart Study. Ethn Dis 2005;15:S6-30-7.

34. Bild DE, Bluemke DA, Burke GL, et al. Multi-Ethnic Study of Atherosclerosis: Objectives and Design. American Journal of Epidemiology 2002;156:871–81.

35. Anderson GL, Manson J, Wallace R, et al. Implementation of the Women’s Health Initiative study design. Ann Epidemiol 2003;13:S5–17.

36. Taliun D, Harris DN, Kessler MD, et al. Sequencing of 53,831 diverse genomes from the NHLBI TOPMed Program. Nature 2021;590:290–9.

37. Ding J, Sidore C, Butler TJ, et al. Assessing Mitochondrial DNA Variation and Copy Number in Lymphocytes of ∼2,000 Sardinians Using Tailored Sequencing Analysis Tools. PLoS Genet 2015;11:e1005306.

38. Barbalic M, Reiner AP, Wu C, et al. Genome-wide association analysis of incident coronary heart disease (CHD) in African Americans: a short report. PLoS Genet 2011;7:e1002199.

39. Adams HP, Jr., Bendixen BH, Kappelle LJ, et al. Classification of subtype of acute ischemic stroke. Definitions for use in a multicenter clinical trial. TOAST. Trial of Org 10172 in Acute Stroke Treatment. Stroke 1993;24:35–41.

40. Mudd JO, Kass DA. Tackling heart failure in the twenty-first century. Nature 2008;451:919–28.

41. Rosamond WD, Chang PP, Baggett C, et al. Classification of heart failure in the atherosclerosis risk in communities (ARIC) study: a comparison of diagnostic criteria. Circ Heart Fail 2012;5:152–9.

42. Bruce Alberts AJ, Julian Lewis, Martin Raff, Keith Roberts, Peter Walter. Molecular Biology of the Cell, 4th edition. New York: Garland Science 2002;ISBN-10: 0-8153-3218-1.

43. Voet DVJ, Pratt CW. Fundamentals of Biochemistry. 2nd Edition. John Wiley and Sons, Inc. 2005:pp. 547. ISBN 0471214957..

44. Hemani G, Zheng J, Elsworth B, et al. The MR-Base platform supports systematic causal inference across the human phenome. Elife 2018;7.

45. Bennet AM, Reynolds CA, Gatz M, Blennow K, Pedersen NL, Prince JA. Pleiotropy in the presence of allelic heterogeneity: alternative genetic models for the influence of APOE on serum LDL, CSF amyloid-beta42, and dementia. J Alzheimers Dis 2010;22:129–34.

46. Verbanck M, Chen C-Y, Neale B, Do R. Detection of widespread horizontal pleiotropy in causal relationships inferred from Mendelian randomization between complex traits and diseases. Nature Genetics 2018;50:693–8.

47. Walker V, Davies N, Hemani G, et al. Using the MR-Base platform to investigate risk factors and drug targets for thousands of phenotypes [version 2; peer review: 3 approved]. Wellcome Open Research 2019;4.

48. Burgess S, Thompson SG. Interpreting findings from Mendelian randomization using the MR-Egger method. Eur J Epidemiol 2017;32:377–89.

49. Bowden J, Davey Smith G, Burgess S. Mendelian randomization with invalid instruments: effect estimation and bias detection through Egger regression. Int J Epidemiol 2015;44:512–25.

50. Bowden J, Davey Smith G, Haycock PC, Burgess S. Consistent Estimation in Mendelian Randomization with Some Invalid Instruments Using a Weighted Median Estimator. Genet Epidemiol 2016;40:304–14.

51. Hartwig FP, Davey Smith G, Bowden J. Robust inference in summary data Mendelian randomization via the zero modal pleiotropy assumption. Int J Epidemiol 2017;46:1985–98.

52. Mi H, Muruganujan A, Huang X, et al. Protocol Update for large-scale genome and gene function analysis with the PANTHER classification system (v.14.0). Nat Protoc 2019;14:703–21.

53. Burgess S, Freitag DF, Khan H, Gorman DN, Thompson SG. Using multivariable Mendelian randomization to disentangle the causal effects of lipid fractions. PLoS One 2014;9:e108891.

54. Davies NM, Holmes MV, Davey Smith G. Reading Mendelian randomisation studies: a guide, glossary, and checklist for clinicians. BMJ 2018;362:k601.

55. Sanderson E, Davey Smith G, Windmeijer F, Bowden J. An examination of multivariable Mendelian randomization in the single-sample and two-sample summary data settings. Int J Epidemiol 2019;48:713–27.

56. Burgess S, Thompson DJ, Rees JMB, Day FR, Perry JR, Ong KK. Dissecting Causal Pathways Using Mendelian Randomization with Summarized Genetic Data: Application to Age at Menarche and Risk of Breast Cancer. Genetics 2017;207:481–7.

57. Relton CL, Davey Smith G. Two-step epigenetic Mendelian randomization: a strategy for establishing the causal role of epigenetic processes in pathways to disease. Int J Epidemiol 2012;41:161–76.

58. Rees JMB, Wood AM, Burgess S. Extending the MR-Egger method for multivariable Mendelian randomization to correct for both measured and unmeasured pleiotropy. Stat Med 2017;36:4705–18.

59. Burgess S, Thompson SG. Multivariable Mendelian randomization: the use of pleiotropic genetic variants to estimate causal effects. Am J Epidemiol 2015;181:251–60.

60. Burgess S, Davies NM, Thompson SG. Bias due to participant overlap in two-sample Mendelian randomization. Genet Epidemiol 2016;40:597–608.

61. Longchamps RJ, Yang SY, Castellani CA, et al. Genome-wide analysis of mitochondrial DNA copy number reveals loci implicated in nucleotide metabolism, platelet activation, and megakaryocyte proliferation. Hum Genet 2022;141:127–46.

62. Lyon M, Andrews SJ, Elsworth B, Gaunt TR, Hemani G, Marcora E. The variant call format provides efficient and robust storage of GWAS summary statistics. bioRxiv 2020:2020.05.29.115824.

63. Elsworth B, Lyon M, Alexander T, et al. The MRC IEU OpenGWAS data infrastructure. bioRxiv 2020:2020.08.10.244293.

64. Nelson CP, Goel A, Butterworth AS, et al. Association analyses based on false discovery rate implicate new loci for coronary artery disease. Nat Genet 2017;49:1385–91.

65. Kanoni S, Graham SE, Wang Y, et al. Implicating genes, pleiotropy and sexual dimorphism at blood lipid loci through multi-ancestry meta-analysis. medRxiv 2021:2021.12.15.21267852.

66. Ramdas S, Judd J, Graham SE, et al. A multi-layer functional genomic analysis to understand noncoding genetic variation in lipids. bioRxiv 2021:2021.12.07.470215.

67. Baigent C, Keech A, Kearney PM, et al. Efficacy and safety of cholesterol-lowering treatment: prospective meta-analysis of data from 90,056 participants in 14 randomised trials of statins. Lancet 2005;366:1267–78.

68. Zorov DB, Filburn CR, Klotz LO, Zweier JL, Sollott SJ. Reactive oxygen species (ROS)-induced ROS release: a new phenomenon accompanying induction of the mitochondrial permeability transition in cardiac myocytes. J Exp Med 2000;192:1001–14.

69. Zorov DB, Juhaszova M, Sollott SJ. Mitochondrial reactive oxygen species (ROS) and ROS-induced ROS release. Physiol Rev 2014;94:909–50.

70. Galkina E, Ley K. Immune and inflammatory mechanisms of atherosclerosis (*). Annu Rev Immunol 2009;27:165–97.

71. Davey Smith G, Hemani G. Mendelian randomization: genetic anchors for causal inference in epidemiological studies. Hum Mol Genet 2014;23:R89–R98.

72. Graham SE, Clarke SL, Wu KH, et al. The power of genetic diversity in genome-wide association studies of lipids. Nature 2021;600:675–9.

73. Kanoni S, Graham SE, Wang Y, et al. Implicating genes, pleiotropy and sexual dimorphism at blood lipid loci through multi-ancestry meta-analysis. medRxiv 2021:2021.12.15.21267852.

74. Ramdas S, Judd J, Graham SE, et al. A multi-layer functional genomic analysis to understand noncoding genetic variation in lipids. bioRxiv 2021:2021.12.07.470215.

75. Psaty BM, Delaney JA, Arnold AM, et al. Study of Cardiovascular Health Outcomes in the Era of Claims Data: The Cardiovascular Health Study. Circulation 2016;133:156–64.

76. Tsao CW, Vasan RS. Cohort Profile: The Framingham Heart Study (FHS): overview of milestones in cardiovascular epidemiology. Int J Epidemiol 2015;44:1800–13.

