## Supplemental materials for "The association between mitochondrial DNA copy number, low-density lipoprotein cholesterol, and cardiovascular disease risk"

**Study population**

This study included participants from eight prospective cohort studies described below. The Atherosclerosis Risk in Communities study (ARIC) (n=3,585) is a prospective epidemiologic study that recruited participants from four communities including Forsyth County, NC; Jackson, MS; the northwest suburbs of Minneapolis, MN; and Washington County, MD. The ARIC analyses focused on CVD outcomes that were adjudicated through 2017 by expert committee that reviewed death certificates, hospital records and telephone interviews. Genomic DNA was extracted in buffy coat from whole blood using the Gentra Puregene Blood Kit (Qiagen) at several health exam visits. mtDNA CN was available for 2,964 participants of European Americans and African Americans with WGS from TOPMed.

Cohort acknowledgement/support: The Atherosclerosis Risk in Communities study has been funded in whole or in part with Federal funds from the National Heart, Lung, and Blood Institute, National Institutes of Health, Department of Health and Human Services, under Contract nos. (75N92022D00001, 75N92022D00002, 75N92022D00003, 75N92022D00004, 75N92022D00005). The authors thank the staff and participants of the ARIC study for their important contributions. WGS for “NHLBI TOPMed: Atherosclerosis Risk in Communities” (phs001211.v3.p2.c1) was performed at the Baylor College of Medicine Human Genome Sequencing Center (3U54HG003273-12S2 / HHSN268201500015C). Core support including centralized genomic read mapping and genotype calling, along with variant quality metrics and filtering were provided by the TOPMed Informatics Research Center (3R01HL-117626-02S1; contract HHSN268201800002I). Core support including phenotype harmonization, data management, sample-identity QC, and general program coordination were provided by the TOPMed Data Coordinating Center (R01HL-120393; U01HL-120393; contract HHSN268201800001I). We gratefully acknowledge the study participants who provided biological samples and data for TOPMed.

The Coronary Artery Risk Development in Young Adults Study (CARDIA) (n=3,411) is a prospective cohort study. Initiated in 1985-86, the CARDIA was aimed to investigate lifestyle and other factors that influence CVD during young adulthood. The study recruited and examined 5,115 Black and White women and men aged 18-30 years in four urban areas: Birmingham, Alabama; Chicago, Illinois; Minneapolis, Minnesota, and Oakland, California. The initial examination included carefully standardized measurements of major risk factors for CVD such as blood pressure, cholesterol and other lipids, and glucose. Data have also been collected on physical measurements such as weight and body composition as well as lifestyle factors. mtDNA CN was available for 3,452 participants with WGS sequencing in TOPMed.

Cohort acknowledgement/support: Cohort acknowledgement/support: WGS for the NHLBI TOPMed program: CARDIA (phs001612) was performed at the Baylor Human Genome Sequencing Center (HHSN268201600033I). Centralized read mapping and genotype calling, along with variant quality metrics and filtering were provided by the TOPMed Informatics Research Center (3R01HL-117626-02S1). Phenotype harmonization, data management, sample-identity QC, and general study coordination, were provided by the TOPMed Data Coordinating Center (3R01HL-120393-02S1). The CARDIA study is conducted and supported by the National Heart, Lung, and Blood Institute (NHLBI) in collaboration with the University of Alabama at Birmingham (HHSN268201800005I & HHSN268201800007I), Northwestern University (HHSN268201800003I), University of Minnesota (HHSN268201800006I), and Kaiser Foundation Research Institute (HHSN268201800004I). We gratefully acknowledge the study participants who provided biological samples and data for TOPMed. We also wish to thank the staff of the CARDIA study.

The Cardiovascular Health Study (CHS) (n=3,539) is a population based, longitudinal, multicenter study of coronary heart disease and stroke in 5,888 elderly adults aged 65 years and older. The CHS originated in 1988 to recruit participants from four U.S. communities. The original cohort recruited 5,201 participants and 687 predominately African-American participants were recruited at three of the four field centers in 1992. The first exam began in June 1989, followed by the second exam 3 years later. A total of n=3,493 CHS participants (mean age 74 and 58% women) with WGS were included in this study.

Cohort acknowledgement/support: Cardiovascular Health Study: This research was supported by contracts HHSN268201200036C, HHSN268200800007C, HHSN268201800001C, N01HC55222, N01HC85079, N01HC85080, N01HC85081, N01HC85082, N01HC85083, N01HC85086, 75N92021D00006, and grants U01HL080295, U01HL130114, and R01HL105756 from the National Heart, Lung, and Blood Institute (NHLBI), with additional contribution from the National Institute of Neurological Disorders and Stroke (NINDS). Additional support was provided by R01AG023629 from the National Institute on Aging (NIA). A full list of principal CHS investigators and institutions can be found at CHS-NHLBI.org. The content is solely the responsibility of the authors and does not necessarily represent the official views of the National Institutes of Health. Sequencing was supported and conducted in collaboration with Baylor University (HHSN268201600033I, 3U54HG003273-12S2, HHSN268201500015C) and Broad Genomics (**HHSN268201600034I)** contracts from NHLBI.

The Framingham Heart Study (FHS) (n=4,133) is a single-site, community-based, prospective study that was initiated in 1948 to investigate the risk factors for CVD. The second generation and the third generation was recruited in 1971 and 2002, respectively. A small number of spouse individuals of the second generation was examined at the same time when the third generation had their first examination. The first generation has been examined every two years. The second and the third generations have been examined every 4-8 years. A total of 4,196 FHS participants were whole genome sequenced by TOPMed. This study included 4,133 FHS participants.

Cohort acknowledgement/support: The WGS for FHS (phs000974) was performed at the Broad Institute of MIT and Harvard (3R01HL092577-06S1 and 3U54HG003067-12S2). The FHS acknowledges the support of contracts NO1-HC-25195, HHSN268201500001I and 75N92019D00031 from the National Heart, Lung and Blood Institute and grant supplement R01 HL092577-06S1 for this research. We also acknowledge the dedication of the FHS study participants without whom this research would not be possible. Dr. Vasan is supported in part by the Evans Medical Foundation and the Jay and Louis Coffman Endowment from the Department of Medicine, Boston University School of Medicine. X.L., S.S., C.L.S, and C.L. are also supported by R01AG059727. C.L.S and S.S are also supported by AG052409, AG054076 and AG059421.

The Genetic Epidemiology Network of Arteriopathy (GENOA) (n=1,249) enrolled sibships in which at least 2 siblings had essential hypertension diagnosed prior to age 60 years. The initial exam enrolled 1583 non-Hispanic white Americans from Rochester, Minnesota, and 1854 African Americans from Jackson, Mississippi during 1995 to 2000. The second exam re-recruited 80% of participants from 2000 to 2005. The GENOA data consists of biological samples (DNA, serum, urine) as well as demographic, anthropometric, environmental, clinical, biochemical, physiological, and genetic data for understanding the genetic predictors of diseases of the heart, brain, kidney, and peripheral arteries. This study included 1,234 participants of African Americans.

Cohort acknowledgement/support: Support for GENOA was provided by the National Heart, Lung and Blood Institute (HL054457, HL054464, HL054481, HL141292, HL119443, and HL087660) of the National Institutes of Health. Sequencing for the GENOA (phs001345.v1.p1) was performed by the University of Washington Northwest Genomics Center (3R01HL055673-18S1 from the NHLBI and at the Broad Institute of MIT and Harvard (HHSN268201500014C)). The authors also wish to thank the staff and participants of GENOA.

The Jackson Heart Study (JHS) (n=3,406 with WGS, n=5,306 total) is one of the largest prospective, epidemiologic investigation of CVD among African Americans residing in the three counties (Hinds, Madison, and Rankin) that make up the Jackson, Mississippi metropolitan area. Data and biologic materials have been collected from 5,306 participants, including a nested family cohort of 1,498 members of 264 families. The age at enrollment for the unrelated cohort was 35-84 years; the family cohort included related individuals >21 years old. Participants provided extensive medical and social history and had an array of physical and biochemical measurements and diagnostic procedures during a baseline examination (2000-2004), two follow-up examinations (2005-2008 and 2009-2012), and ancillary studies. Samples for genomic DNA were collected during the first two examinations. Consent for genetic studies and broad sharing of genetic data was provided by 3,482 participants. After all quality control procedures, whole genome sequence data are available for 3,406 participants. Follow-up information on vital status, major illnesses or injuries, and hospitalizations to identify intervening clinical events is done annually by phone. Medical records of cardiovascular disease related hospitalizations and death certificates are abstracted and used for adjudication of cardiovascular events and related deaths. Note that heart failure adjudication in JHS only started in 2005 with questions at the annual follow up questionnaire (heart failure case/control status wasn't asked about at baseline visit).

Cohort acknowledgement/support: The Jackson Heart Study (JHS) is supported and conducted in collaboration with Jackson State University (HHSN268201800013I), Tougaloo College (HHSN268201800014I), the Mississippi State Department of Health (HHSN268201800015I) and the University of Mississippi Medical Center (HHSN268201800010I, HHSN268201800011I and HHSN268201800012I) contracts from the National Heart, Lung, and Blood Institute (NHLBI) and the National Institute on Minority Health and Health Disparities (NIMHD). The authors also wish to thank the staffs and participants of the JHS. Molecular data for the Trans-Omics in Precision Medicine (TOPMed) program was supported by the National Heart, Lung and Blood Institute (NHLBI). Genome sequencing for “NHLBI TOPMed: The Jackson Heart Study” (phs000964.v1.p1) was performed at the Northwest Genomics Center (HHSN268201100037C). Core support including centralized genomic read mapping and genotype calling, along with variant quality metrics and filtering were provided by the TOPMed Informatics Research Center (3R01HL-117626-02S1; contract HHSN268201800002I). Core support including phenotype harmonization, data management, sample-identity QC, and general program coordination were provided by the TOPMed Data Coordinating Center (R01HL-120393; U01HL-120393; contract HHSN268201800001I). Laura Raffield was also supported by the National Center for Advancing Translational Sciences, National Institutes of Health, through Grant KL2TR002490 (LMR). We gratefully acknowledge the studies and participants who provided biological samples and data for TOPMed.

**JHS disclaimer-** The views expressed in this manuscript are those of the authors and do not necessarily represent the views of the National Heart, Lung, and Blood Institute; the National Institutes of Health; or the U.S. Department of Health and Human Services.

The Multi-Ethnic Study of Atherosclerosis Study (MESA) (n=4,593) is a study of the characteristics of subclinical cardiovascular disease and the risk factors that predict progression to clinically overt cardiovascular disease or progression of the subclinical disease.^27^ The MESA researchers study a diverse, population-based sample of 6,814 men and women 45-84 years of age and free of prevalent clinical CVD when recruited from six field centers across the United States in 2000-2002. CVD event was adjudicated through 2015 by expert committee to review of death certificates, hospital records. DNA for mtDNA-CN analyses was isolated from exam 1 peripheral leukocytes using the Gentra Puregene Blood Kit. mtDNA CN was available for 4,596 individuals (24.1% Black, 22.3% Hispanic, 13.1% Chinese, and 40.5% White) derived from TOPMed WGS sequencing.

Cohort acknowledgement/support: Whole genome sequencing (WGS) for the Trans-Omics in Precision Medicine (TOPMed) program was supported by the National Heart, Lung and Blood Institute (NHLBI). WGS for “NHLBI TOPMed: Multi-Ethnic Study of Atherosclerosis (MESA)” (phs001416.v1.p1) was performed at the Broad Institute of MIT and Harvard (3U54HG003067-13S1). Centralized read mapping and genotype calling, along with variant quality metrics and filtering were provided by the TOPMed Informatics Research Center (3R01HL-117626-02S1). Phenotype harmonization, data management, sample-identity QC, and general study coordination, were provided by the TOPMed Data Coordinating Center (3R01HL-120393-02S1), and TOPMed MESA Multi-Omics (HHSN2682015000031/HSN26800004). The MESA projects are conducted and supported by the National Heart, Lung, and Blood Institute (NHLBI) in collaboration with MESA investigators. Support for the Multi-Ethnic Study of Atherosclerosis (MESA) projects are conducted and supported by the National Heart, Lung, and Blood Institute (NHLBI) in collaboration with MESA investigators. Support for MESA is provided by contracts 75N92020D00001, HHSN268201500003I, N01-HC-95159, 75N92020D00005, N01-HC-95160, 75N92020D00002, N01-HC-95161, 75N92020D00003, N01-HC-95162, 75N92020D00006, N01-HC-95163, 75N92020D00004, N01-HC-95164, 75N92020D00007, N01-HC-95165, N01-HC-95166, N01-HC-95167, N01-HC-95168, N01-HC-95169, UL1-TR-000040, UL1-TR-001079, UL1-TR-001420, UL1TR001881, DK063491, and R01HL105756. The authors thank the other investigators, the staff, and the participants of the MESA study for their valuable contributions.  A full list of participating MESA investigators and institutes can be found at [http://www.mesa-nhlbi.org](https://nam02.safelinks.protection.outlook.com/?url=http%3A%2F%2Fwww.mesa-nhlbi.org%2F&data=04%7C01%7Cwpost%40jhmi.edu%7Ce250d1c265a847ec090f08d9fbc439a1%7C9fa4f438b1e6473b803f86f8aedf0dec%7C0%7C0%7C637817642593854966%7CUnknown%7CTWFpbGZsb3d8eyJWIjoiMC4wLjAwMDAiLCJQIjoiV2luMzIiLCJBTiI6Ik1haWwiLCJXVCI6Mn0%3D%7C3000&sdata=tVqOVyqCZnioV87T5M39EUUQjtfxVyF4i%2FhQtZTLbwY%3D&reserved=0).

The Women’s Health initiative Study (WHI) (n=7,197) was initiated in 1992, which is a large and complex clinical investigation of strategies for the prevention and control of some of the most common causes of morbidity and mortality among postmenopausal women.^28^ The WHI enrolled women ranging in age from 50 to 79 at one of 40 WHI clinical centers nationwide. The women participants were enrolled into either a clinical trial or observation study. Several of the TOPMed cohorts contained a small number of duplicated participants. After removing the duplicates, This study included up to 27,316 participants (mean age 62, age range of 19-98 years, and 68% women) with 16636 (60.9%) European Americans, 8709 (31.9%) African Americans, 1229 (4.5%) Hispanic/Latino Americans and 728 (2.7%) Chinese Americans (see **Supplemental Table 1**).

Cohort acknowledgement/support: The WHI program is funded by the National Heart, Lung, and Blood Institute, National Institutes of Health, U.S. Department of Health and Human Services through contracts 75N92021D00001, 75N92021D00002, 75N92021D00003, 75N92021D00004, 75N92021D00005. We thank all WHI participants for their dedications and contributions.

**Supplemental Methods**

*WHI study design and weighted cox regression model*

WHI consists of an observational study (n=93k) and three clinical trials (Hormone Therapy=28K, Dietary Modification=48K, Calcium Vit D sups=36K, total 68K). In our study, we included about 11,000 participants who have genetic data in TOPMed. Among these participants, about 5000 of them have incident strokes, 1000 of them have VTE cases and 5000 of them are controls. Most clinical outcomes were adjudicated and outcomes at baseline were collected by self-report.

Given the case-control study design of WHI cohort, we applied weighted cox regression model to analyze the data. The models were as follows:

Model1:

Coxph (Surv(followup time, CVD outcome) ~ mtDNA_stresid+age_blood_draw, weights=whi_weights)

Model2:

Coxph (Surv(followup time, CVD outcome) ~ mtDNA_stresid+age_blood_draw +bmi+tchol+hdlc+sbp+smokenow+baseline_hypertension+baseline_treated_t2d, weights=whi_weights)

Model3:

Coxph (Surv(followup time, CVD outcome) ~ mtDNA_stresid+age_blood_draw +bmi+tchol+hdlc+sbp+smokenow+baseline_hypertension+baseline_treated_t2d+wbc+platelet, weights=whi_weights)

**Supplemental Results**

**Tables (in Excel file)**

**Supplemental Table 1**. Cohort characteristics

**Supplemental Table 2**. Mitochondrial functions list

**Supplemental Table 3**. Stratified analyses

**Supplemental Table 4**. Mendelian Randomization Analysis: The 77 SNPs as instrument variables to infer the causal relationship from mtDNA CN to CHD

**Supplemental Table 5**. Mendelian Randomization Analysis: The 23 SNPs as instrument variables to test the causal relationship from mtDNA CN to CHD

**Supplemental Table 6**. Mendelian analyses: Inverse Variance Weighted analysis and sensitivity analyses

**Supplemental Table 7**. Mendelian Randomization Analysis: The 10 SNPs as instrument variables to test the causal relationship from CHD to mtDNA CN

**Supplemental Table 8**. Mendelian Randomization Analysis: The 75 SNPs as instrument variables to test the causal relationship from mtDNA CN to LDL

**Supplemental Table 9**. Mendelian analyses: Inverse Variance Weighted analysis and sensitivity analyses

**Supplemental Table 10**. Mendelian Randomization Analysis: The 345 SNPs as instrument variables to test the causal relationship from LDL to mtDNA CN

**Figures**

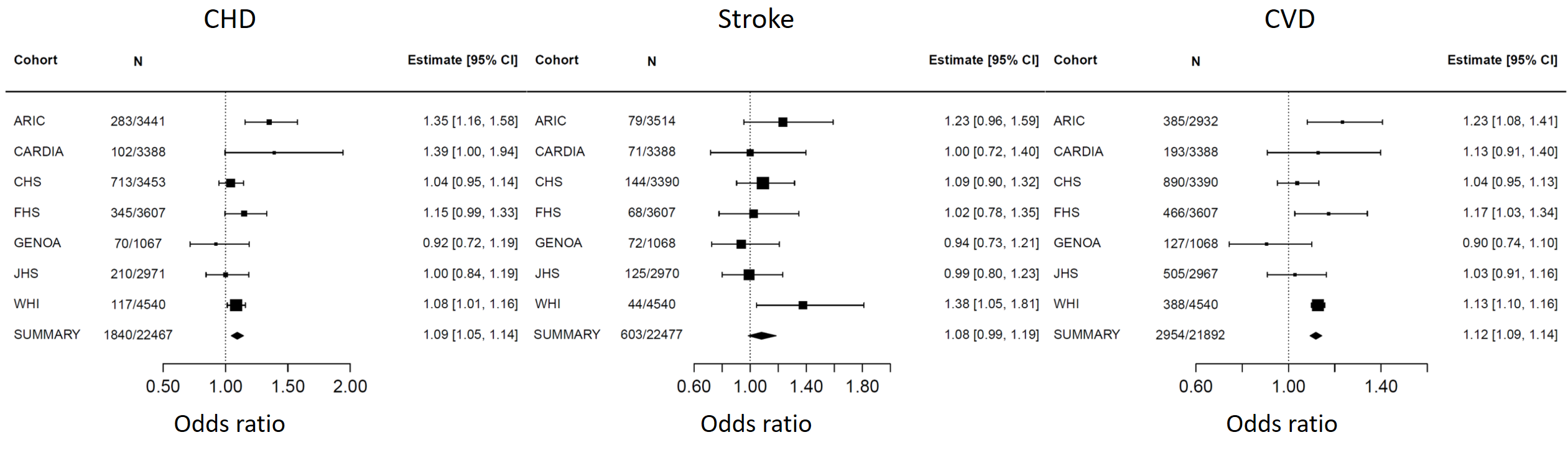

**Supplemental Figure 1.** Association and meta-analysis of mtDNA CN and prevalent CVD outcomes. We performed logistic regression with an outcome and mtDNA residuals as independent variable adjusting for age, sex, study center (if applicable), race/ethnicity, BMI, TC, HDL, SBP, HRX, current smoking and diabetes in Model 2.

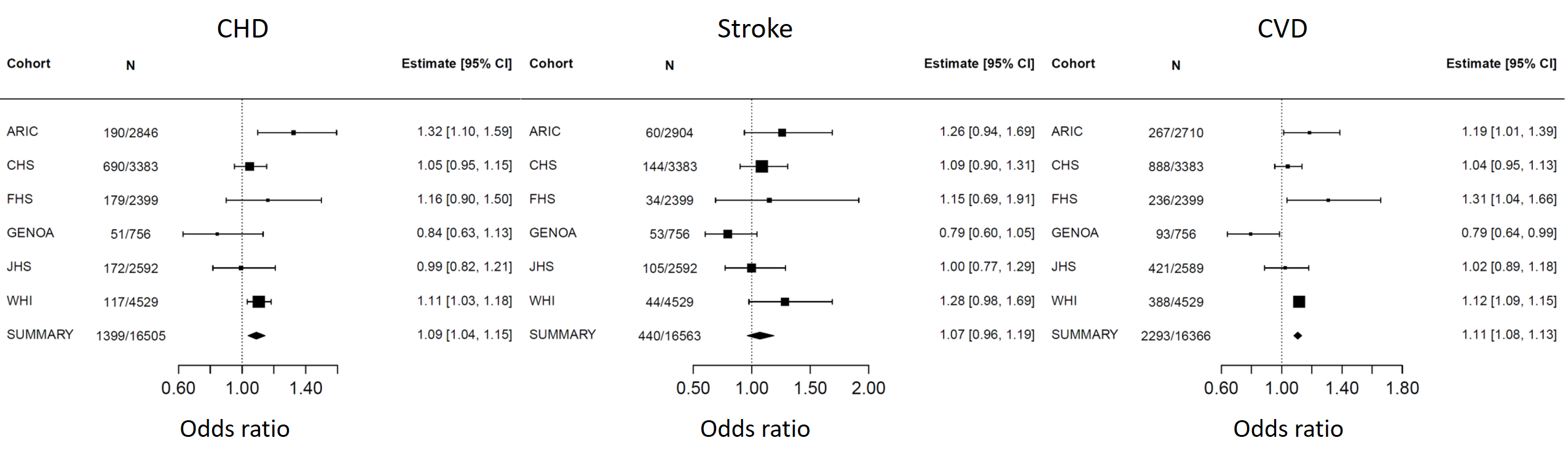

**Supplemental Figure 2.** Association and meta-analysis of mtDNA CN and prevalent CVD outcomes. We performed logistic regression with an outcome and mtDNA residuals as independent variable adjusting for age, sex, study center (if applicable), race/ethnicity, BMI, TC, HDL, SBP, HRX, current smoking, diabetes, WBC, NE and PLT in Model 3.

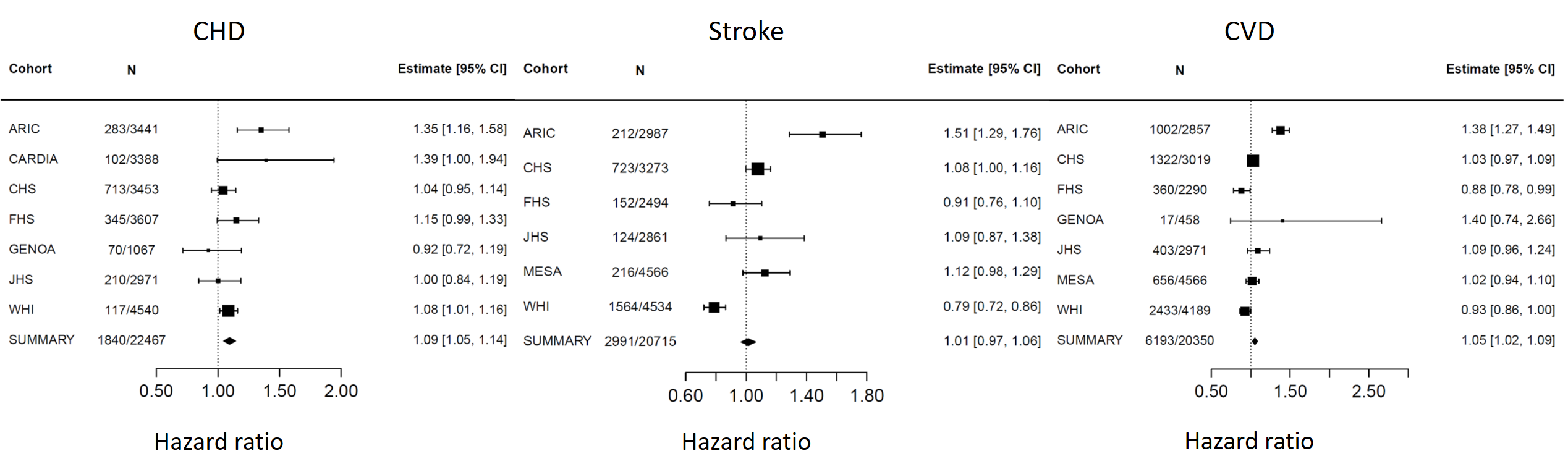

**Supplemental Figure 3**. Association and meta-analysis of mtDNA CN and incident CVD outcomes. We performed cox proportional hazard regression with an outcome and mtDNA residuals as independent variable adjusting for age, sex, study center (if applicable), race/ethnicity, BMI, TC, HDL, SBP, HRX, current smoking and diabetes in Model 2.

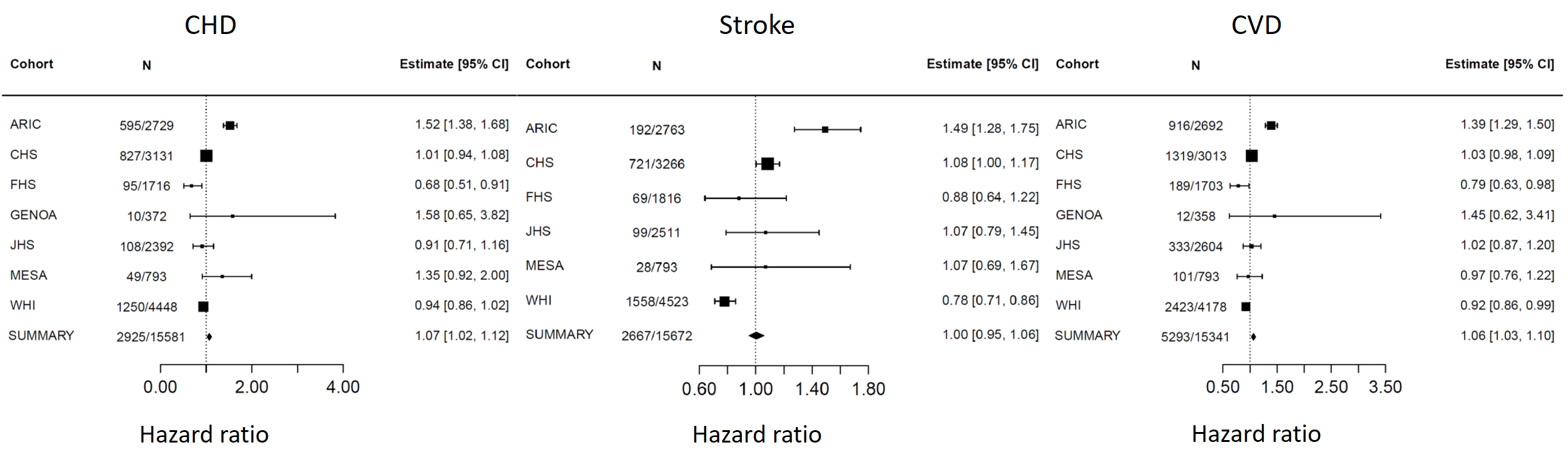

**Supplemental Figure 4**. Association and meta-analysis of mtDNA CN and incident CVD outcomes. We performed cox proportional hazard regression with an outcome and mtDNA residuals as independent variable adjusting for age, sex, study center (if applicable), race/ethnicity, BMI, TC, HDL, SBP, HRX, current smoking, diabetes, WBC, NE and PLT in Model 3.

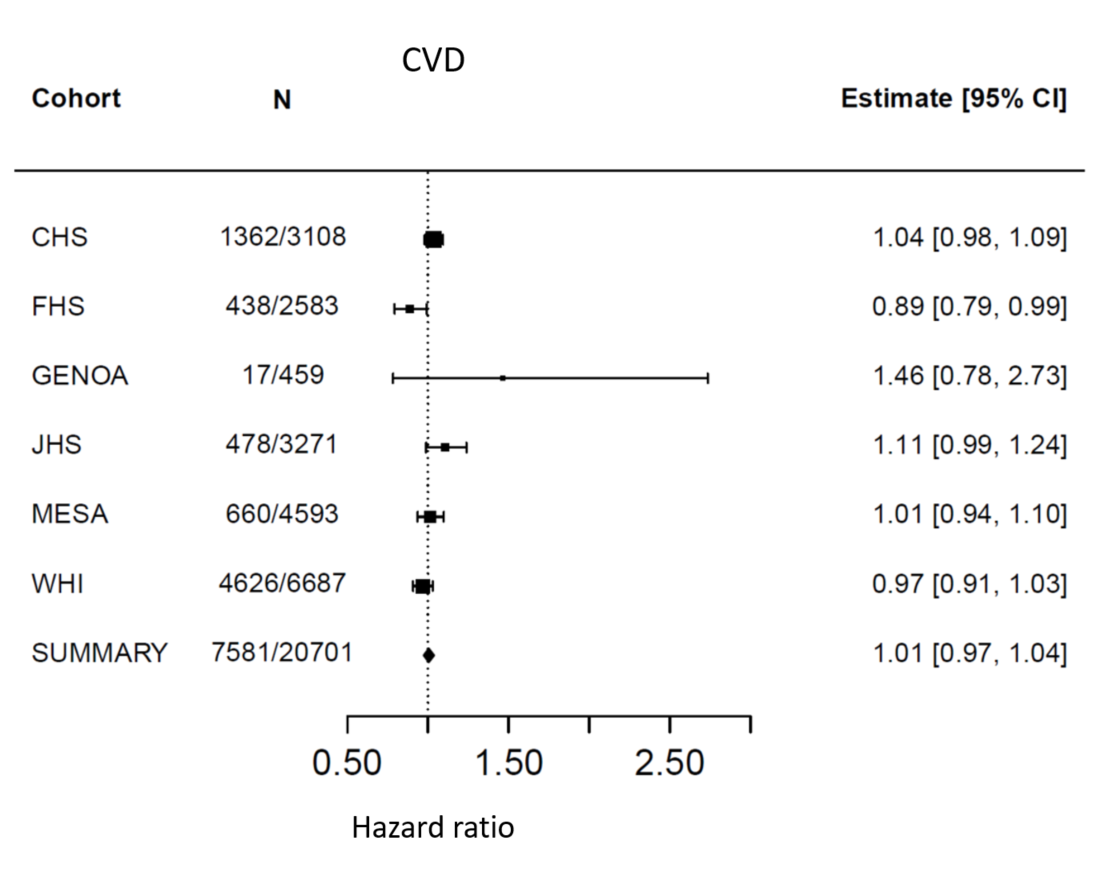

**Supplemental Figure 5**. Association and meta-analysis of mtDNA CN and incident CVD outcomes without ARIC cohort. We performed Cox proportional hazards regression with an outcome and mtDNA residuals as independent variable adjusting for age, sex, study center (if applicable), and race/ethnicity in Model 1.

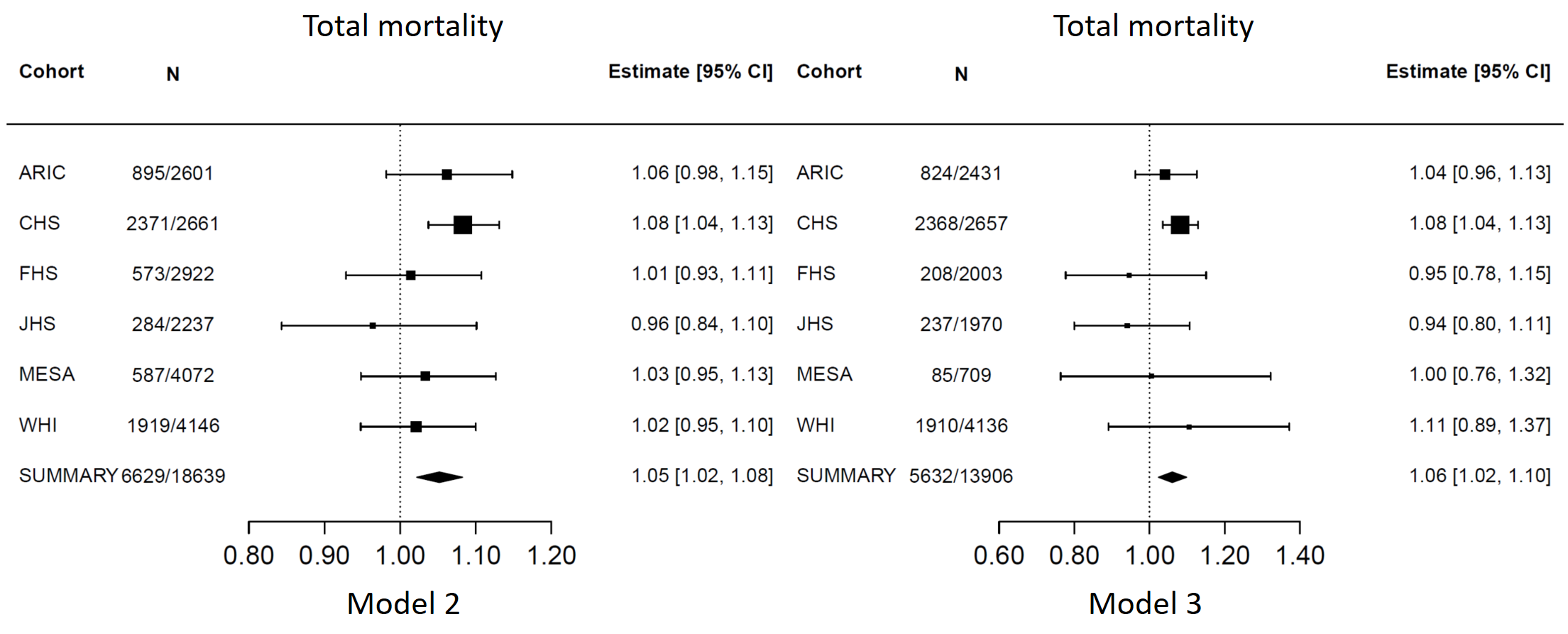

**Supplemental Figure 6**. Association and meta-analysis of mtDNA CN and total mortality. We performed cox proportional hazard regression with an outcome and mtDNA residuals as independent variable adjusting for age, sex, study center (if applicable), race/ethnicity, BMI, TC, HDL, SBP, HRX, current smoking, diabetes, WBC, NE and PLT in Model 2 and further adjusted for WBC, NE and PLT in Model 3.

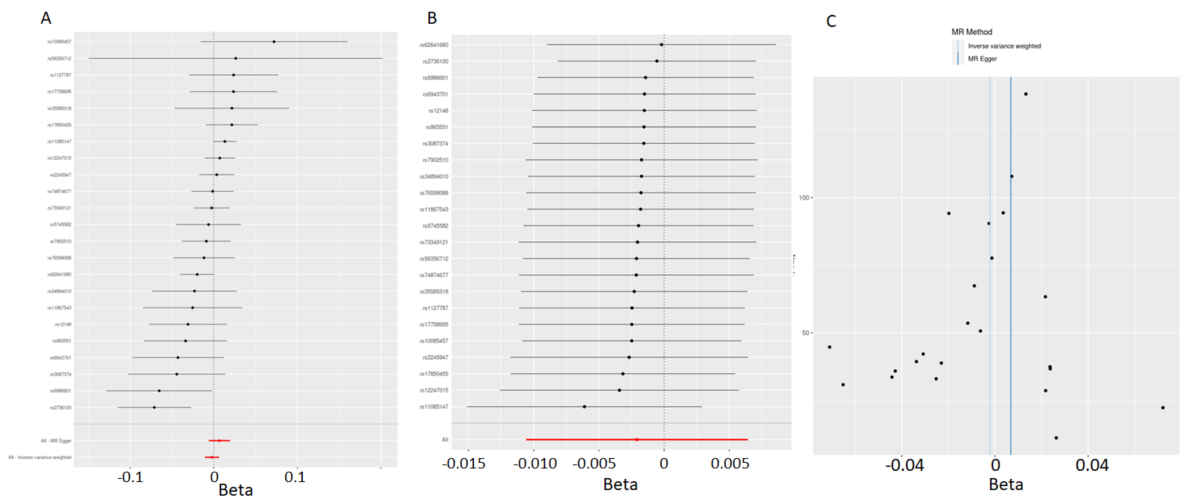

**Supplemental Figure 7.** Forest plot (A), leave one out plot (B) and funnel plot (C) for MR analyses of mtDNA CN on CHD. These SNPs are directly involved in mitochondrial functions.

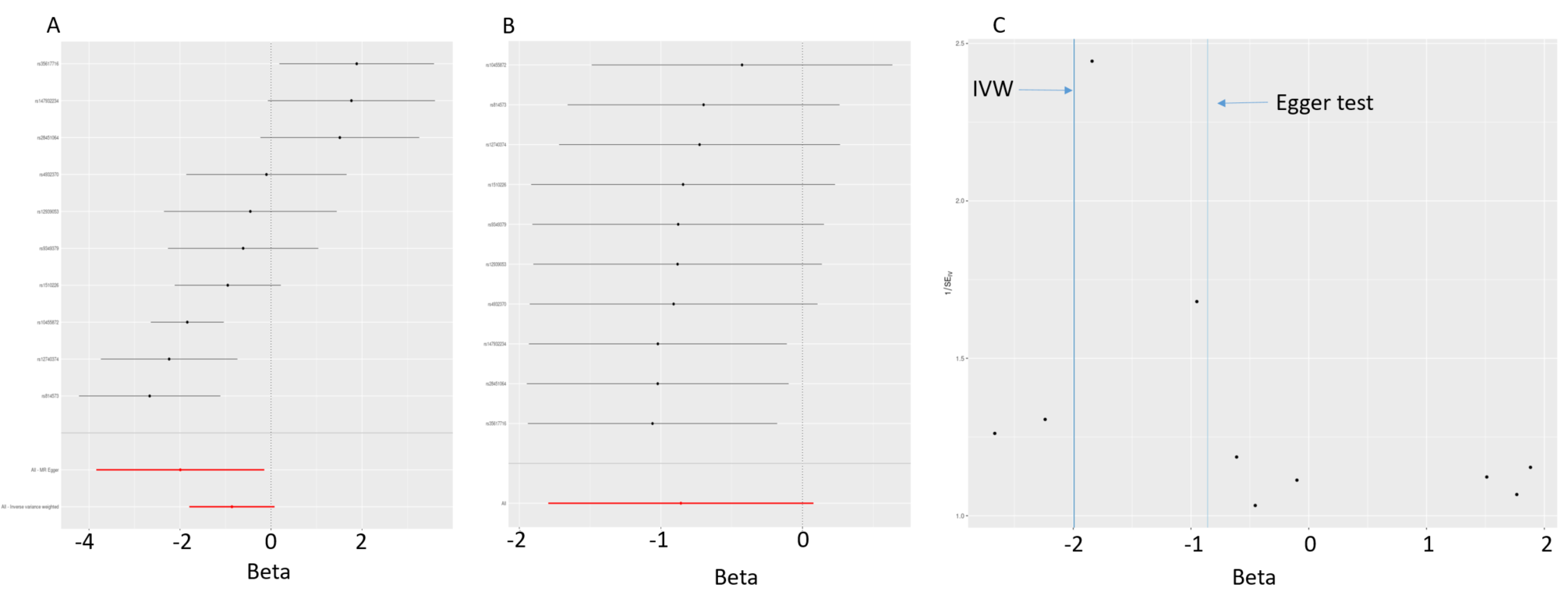

**Supplemental Figure 8.** Forest plot (A), leave one out plot (B) and funnel plot (C) for MR analyses of CHD on mtDNA CN.

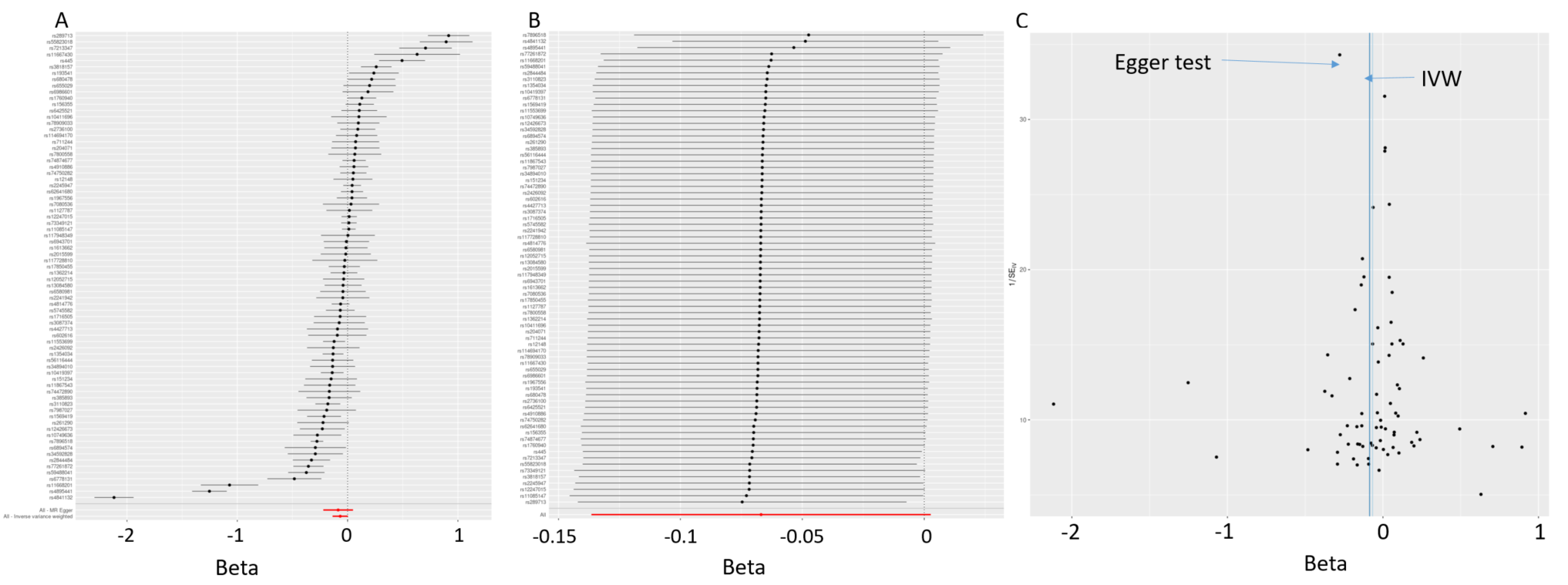

**Supplemental Figure 9.** Forest plot (A), leave one out plot (B) and funnel plot (C) for MR analyses of mtDNA CN on LDL.
